# Endometrial cancer survival disparities in women of African ancestry persist beyond clinical, molecular, and socioeconomic determinants

**DOI:** 10.64898/2026.06.22.26355869

**Authors:** Devin Gee, Alisha Daroch, Meredith Akerman, Natalie Danziger, Leslie Panella, Megan Gorman, Madeline Bright, Douglas I. Lin, Nyasha Chambwe, Marina Frimer

## Abstract

**Introduction:** Stark disparities in endometrial cancer (EC) risk and mortality exist between non-Hispanic Black and White women, with Black women experiencing higher incidence and worse survival. This disparity has been attributed to biological and socioeconomic factors, though how these factors interact to influence EC disparities remains unclear. This study modeled EC outcomes using race, area-level socioeconomic deprivation, clinical phenotypes, genetic ancestry, and molecular alterations.

**Methods:** We identified 281 cases of EC diagnosed from 2013-2023 in women who underwent clinical genomic sequencing as part of routine care across multiple Northwell Health sites. We estimated genetic ancestry, oncogenic alterations in 324 genes, microsatellite instability, and molecular classification. Geocoded patient addresses were used to derive the state-level Area Deprivation Index to estimate socioeconomic deprivation.

**Results:** African ancestry patients were enriched for high-grade disease (89% vs 64%), serous histology (57% vs 26%), and the *TP53-*mutant molecular classification (71% vs 51%) compared to European ancestry patients (p-value<0.05). Socioeconomic deprivation quantiles were associated with race, with more deprived quantiles enriched for Black patients (p-value<0.001). Both race and genetic ancestry, but not area-level deprivation, were independently associated with differences in progression-free survival. *TP53* mutations were enriched in African ancestry patients, while *KRAS*, *PTEN*, and *ARID1A* mutations were enriched in European ancestry patients (q<0.10). Cox proportional hazards modeling, adjusting for these factors, showed that African ancestry patients had worse progression-free survival (HR 1.91, p-value<0.05).

**Conclusion:** Our findings indicate that EC disparities persist after adjusting for socioeconomic, clinical, and molecular factors, highlighting the need to further investigate additional drivers of disparity.

**Highlights:**

- This study modeled drivers of mortality disparities in high-grade endometrial cancer.
- Socioeconomic deprivation was associated with Black or African American race.
- African ancestry was associated with serous, *TP53*-mutated, and higher-grade tumors.
- Features beyond state-level ADI and tumor phenotype may partly explain this gap.

## Introduction

Endometrial cancer (EC) is the most common gynecologic malignancy in the United States and one of the few with increasing mortality, with a 1.5% rise in mortality from 2013 to 2022 [1]. Black women have twofold higher EC mortality compared with White women and women of other racial or ethnic groups [1]. These disparities remain even after adjusting for stage at presentation and receipt of first-line treatment, emphasizing the critical need to understand their underlying drivers [2].

Black women have a 2-4 times higher incidence rate of non-endometrioid EC subtypes [3,4] (including serous carcinoma, clear cell carcinoma, and carcinosarcoma), which represent only 5-10% of EC diagnoses but constitute 40% of EC deaths [4]. Molecular classification using the TCGA classification scheme [5] shows that Black women are disproportionately affected by molecular subtypes characterized by high copy-number variation and frequent *TP53* mutations (TP53mut), which are associated with significantly worse survival [6,7]. Despite these well-established disparities, Black women remain underrepresented in many clinical and molecular research cohorts, limiting comprehensive understanding of tumor biology and further limiting the development of targeted therapies.

Increasing evidence points to an influential role of the patient’s lived environment on EC mortality. For instance, county-level socioeconomic characteristics have been associated with poorer survival rates in women with uterine cancer, especially among women from underrepresented racial and ethnic groups [8]. One composite measure of the lived environment, the Area Deprivation Index (ADI), uses 17 indicators of socioeconomic status to estimate neighborhood disadvantage [9,10]. Higher ADI, thus greater deprivation, has been associated with poorer EC-specific mortality, though disparities in mortality between races remain across the ADI distribution [11,12]. A difference in mortality between races has been observed in equal-access healthcare systems, suggesting multifactorial drivers in EC disparities beyond socioeconomic deprivation or access to care [13].

Race and genetic ancestry represent related but distinct characteristics. Race represents the social and cultural identities that one self-identifies with, while genetic ancestry is a continuous spectrum referring to the populations from which one inherits their alleles [14]. Here we use race and predominant continental genetic ancestry to categorize patients and evaluate differences in survival outcomes.

Though the association between race, ancestry, tumor subtypes, molecular characteristics, and socioeconomic deprivation in EC mortality has been established individually, these factors do not act in isolation. There is a critical need to understand whether the combination of these factors explains the extent of EC disparities and determine if undiscovered drivers remain. This study aims to model the interplay of these biological and socioeconomic factors to determine whether they fully account for EC disparities.

## Methods

### Data Collection

This was a multi-site retrospective study that included patients with a histologic diagnosis of EC who underwent FoundationOne® or FoundationOne® CDx genomic sequencing as part of their routine clinical workflow. IRB approval was obtained at Northwell Health (IRB#: 19-0259-LIJ). Demographic and clinical data were obtained through retrospective chart review using the electronic medical record and study variables were stored and managed using a Research Electronic Data Capture (REDCap) database instance hosted at Northwell Health [15,16]. Study variables captured include primary tumor site, surgical history, pathologic findings including histologic subtype, grade, stage of disease, use of neoadjuvant chemotherapy, standard treatment modalities and duration received, and first recurrence event with subsequent therapies. The date of last follow-up or death was also recorded. The study participants’ primary address at diagnosis was extracted and recorded for further analysis.

### Geocoding and Area Deprivation Index

Study subjects’ primary addresses were geocoded using the Geocodio Application Programming Interface through the tidygeocoder R package to determine longitudes and latitudes. These values were assigned their respective United States census block Federal Information Processing System (FIPS) codes. FIPS codes were used to assign the 2022 ADI national percentile and ADI state decile for each patient census block group [10]. State Deciles were grouped into quantiles, with the last two quantiles (Q4 and Q5) combined due to low sample sizes.

### Comprehensive Genomic Profiling

Comprehensive Genomic Profiling (CGP) was performed using FoundationOne® CDx or its laboratory-developed test predecessor FoundationOne® by hybrid-capture-based next-generation sequencing in a Clinical Laboratory Improvement Amendments (CLIA)-certified, College of American Pathologists (CAP) accredited laboratory (Foundation Medicine, Inc., Boston, MA) as previously described [17,18]. In addition to the evaluation of genomic alterations in up to 324 genes, microsatellite instability (MSI), tumor mutational burden (TMB), and a homologous recombination deficiency signature (HRDsig), provided as a laboratory professional service, were calculated as previously described [19–22].

Homologous repair (HR) alterations were defined as alterations in *BRCA1*, *BRCA2*, *PALB2*, *BRIP1*, *RAD51C*, or *RAD51D*. Genomic loss of heterozygosity (gLOH), provided as a laboratory professional service, was calculated by quantifying LOH at > 3,500 sequenced single nucleotide polymorphisms (SNPs), excluding whole chromosome arm losses (defined as > 90% loss of the arm) and SNPs with ≥ 40%. Patients above 16% were labeled gLOH-High and patients below 16% were labeled gLOH-Low.

### Molecular Classification

Molecular classification was performed using CGP results as previously described [23]. Tumors with pathogenic *POLE* alterations were categorized as POLEmut. Tumors wildtype for *POLE* (POLEwt) with high microsatellite instability (MSI-H) were categorized as MSI-H. Tumors that were POLEwt and microsatellite stable (MSS) with *TP53* alterations were assigned TP53mut. The remaining tumors (POLEwt, MSS, TP53wt) were assigned ‘no specific molecular profile’ (NSMP). Tumors with indeterminate MSI status were labeled as cannot be determined (CBD).

### Genetic Ancestry Estimation

Estimated genetic ancestry for each patient was derived from the CGP assay using a SNP-based approach as previously described [24,25]. Briefly, a random forest classifier was trained using over 40,000 germline SNPs, with cross-validation performed using the 1000 Genomes Project (1KGP) cohort as a reference panel. Predominant genetic ancestry was classified into the following five groups: European (EUR), African (AFR), East Asian (EAS), South Asian (SAS), and admixed American (AMR), corresponding to the major continental groups defined by the 1KGP.

### Statistical Methods

Descriptive statistics (frequencies and percentages for categorical variables) were calculated for the overall sample, by Genetic Ancestry (European vs. African), by Race (White vs. Black or African American, and by ADI quantile). Groups were compared using the chi-square test or Fisher’s exact test, as deemed appropriate. To assess significant differences in gene alteration frequencies between comparison groups we used the chi-square test or Fisher’s exact test.

Clinical endpoints evaluated in this study were progression-free survival (PFS) and overall survival (OS). PFS was defined as the time from the date of diagnosis to the date of documented disease progression or death from any cause, whichever occurs first. OS was defined as the time from the date of diagnosis to the date of death from any cause. If no disease progression or death is documented prior to study termination, these endpoints were censored at the date of last follow-up.

Survival analyses were conducted to evaluate associations between baseline clinical, demographic, and molecular characteristics and both OS and PFS. The Kaplan-Meier method was used to estimate survival distributions, and differences between groups were assessed using the log-rank test. To further quantify associations, Cox proportional hazards regression models were employed. Both univariable and multivariable Cox models were constructed to assess the impact of individual covariates on the outcomes. Covariates included in the models were ancestry, ADI quantile, age, FIGO 2009 stage, tumor grade, histology, molecular classification, tumor mutational burden (TMB), homologous recombination deficiency (HRD) status, and body mass index (BMI). Multivariable models incorporated covariates that were either clinically relevant or statistically significant in univariable analyses.

Proportional hazards assumptions were evaluated using Schoenfeld residuals and graphical diagnostics. Results are reported as hazard ratios (HRs) with corresponding 95% confidence intervals (CIs). A result was considered statistically significant at p < 0.05. All analyses were performed using SAS version 9.4 (SAS Institute Inc., Cary, NC), R version 4.2.1, and RStudio version 2024.09.0+375.

## Results

### Cohort Overview

We reviewed data from 321 uterine cancer cases from patients who underwent genomic sequencing as part of routine clinical care. Patients with non-endometrial carcinoma malignancies at diagnosis or insufficient clinical data were excluded. Patients who could not be geocoded or who listed a primary address outside of New York state were removed from analysis. Tumors with POLEmut molecular classification were removed due to low sample size resulting in 281 eligible patients (**Figure 1**). Most study eligible patients presented with advanced disease, with 164 (58.4%) at stage III or IV and 205 (73.0%) having grade 3 tumors (**Table 1**). High-grade tumors were primarily composed of non-endometrioid subtypes, including serous (47.3%), carcinosarcoma (22.4%), and clear cell histologies (10.7%) (**Table 1**). TP53mut was the predominant molecular classification (51.6%), followed by NSMP (21.4%), MSI-H (19.2%), and CBD (7.8%) (**Table 1**). Patients were racially diverse, with 163 (58.0%) patients identified as White, 75 (26.7%) identified as Black or African American and 33 (11.7%) identified as Asian (**Table 1**). Based on genetic ancestry estimation, 76 (27.0%) patients had predominantly AFR ancestry while 148 (52.7%) had predominantly EUR ancestry (**Table 1**). To best account for the heterogeneity of patients in our cohort, we primarily use genetic ancestry to provide finer categories to correlate with cohort characteristics.

**Figure 1.**
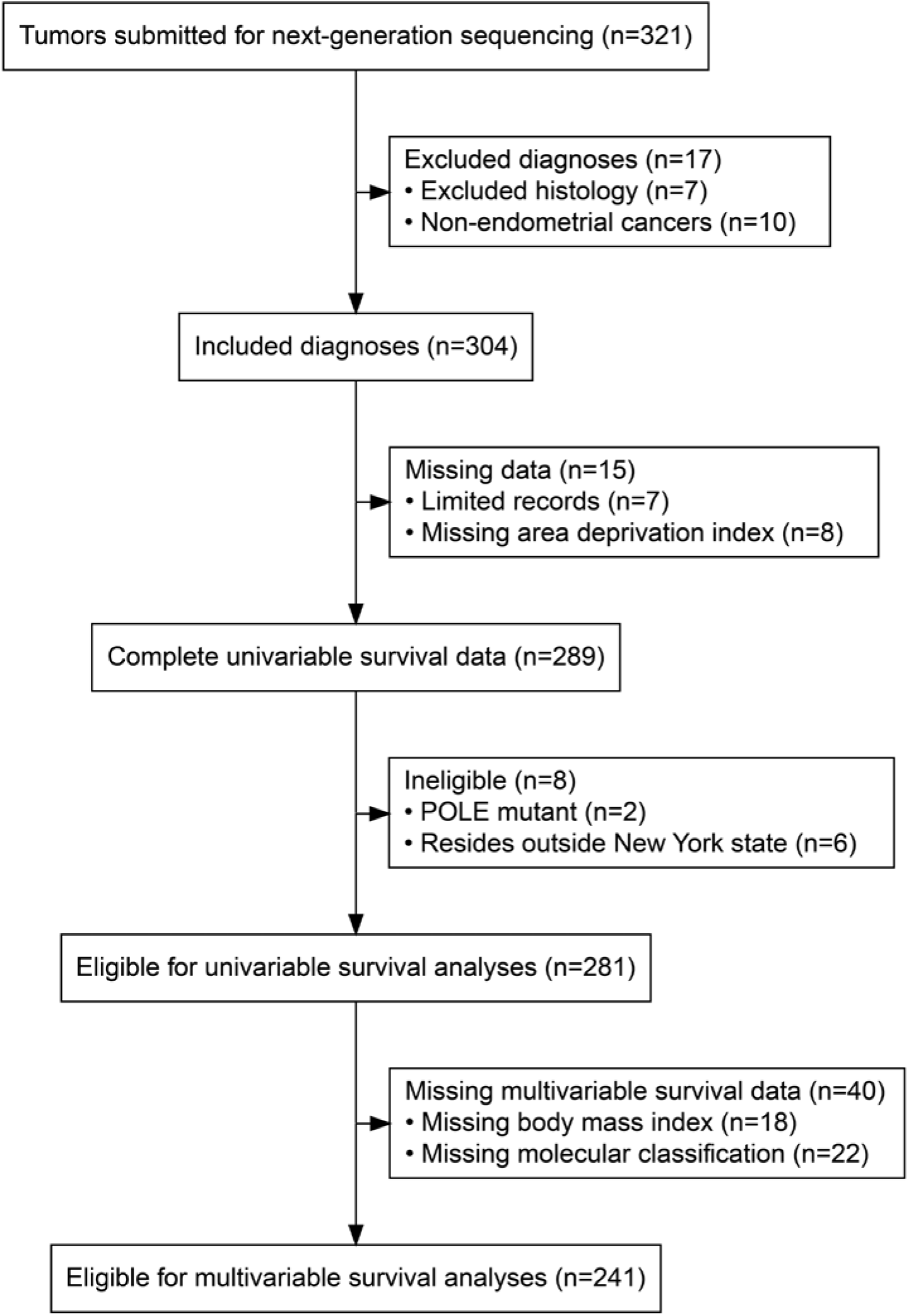
Patient Eligibility Criteria: 281 patients were included for univariable survival analyses, and 241 patients were included for multivariable survival analyses. Exclusions were due to cancer type and histology at diagnosis, data completeness, New York State residence status, and molecular classification.

**Table 1.**
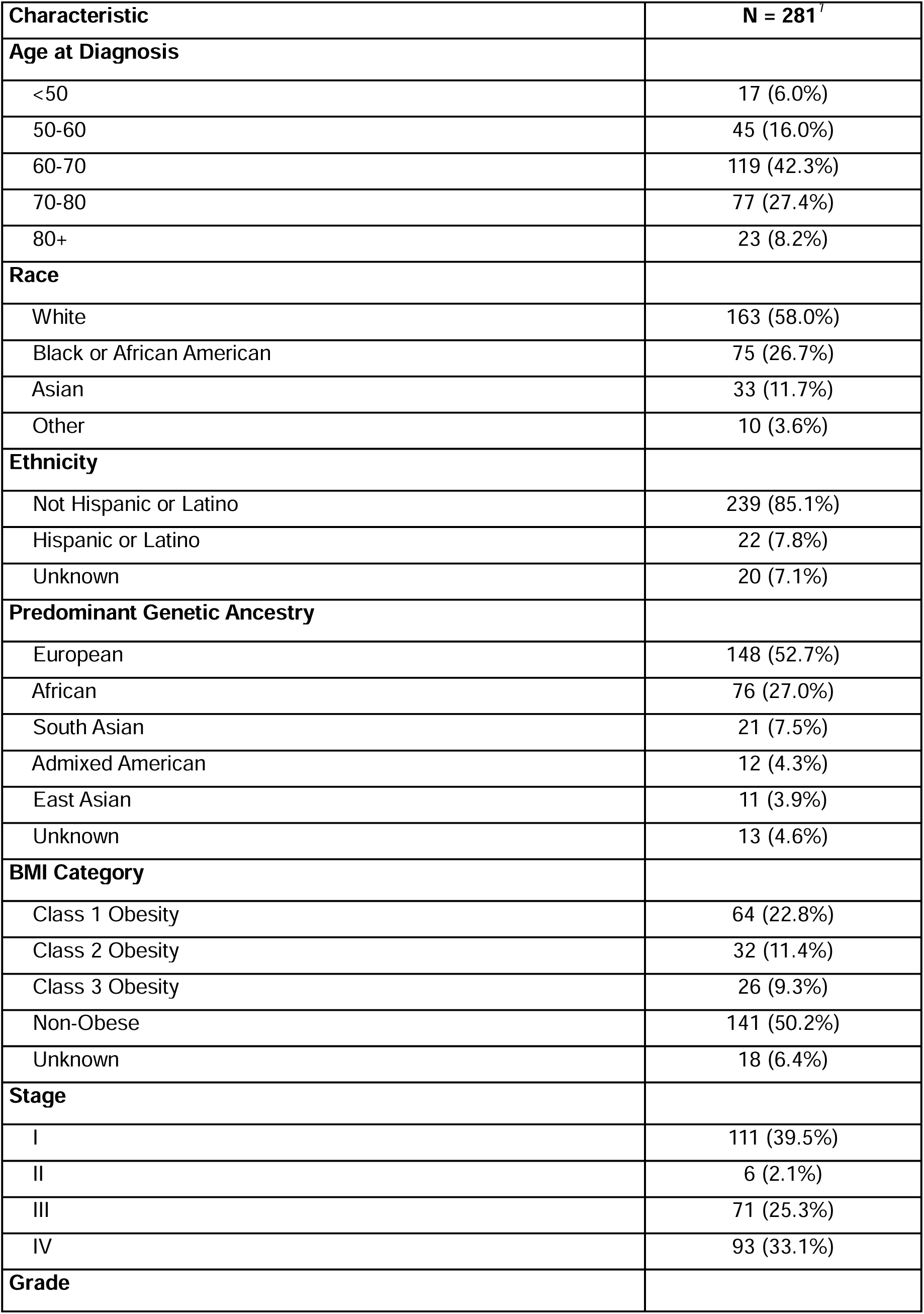
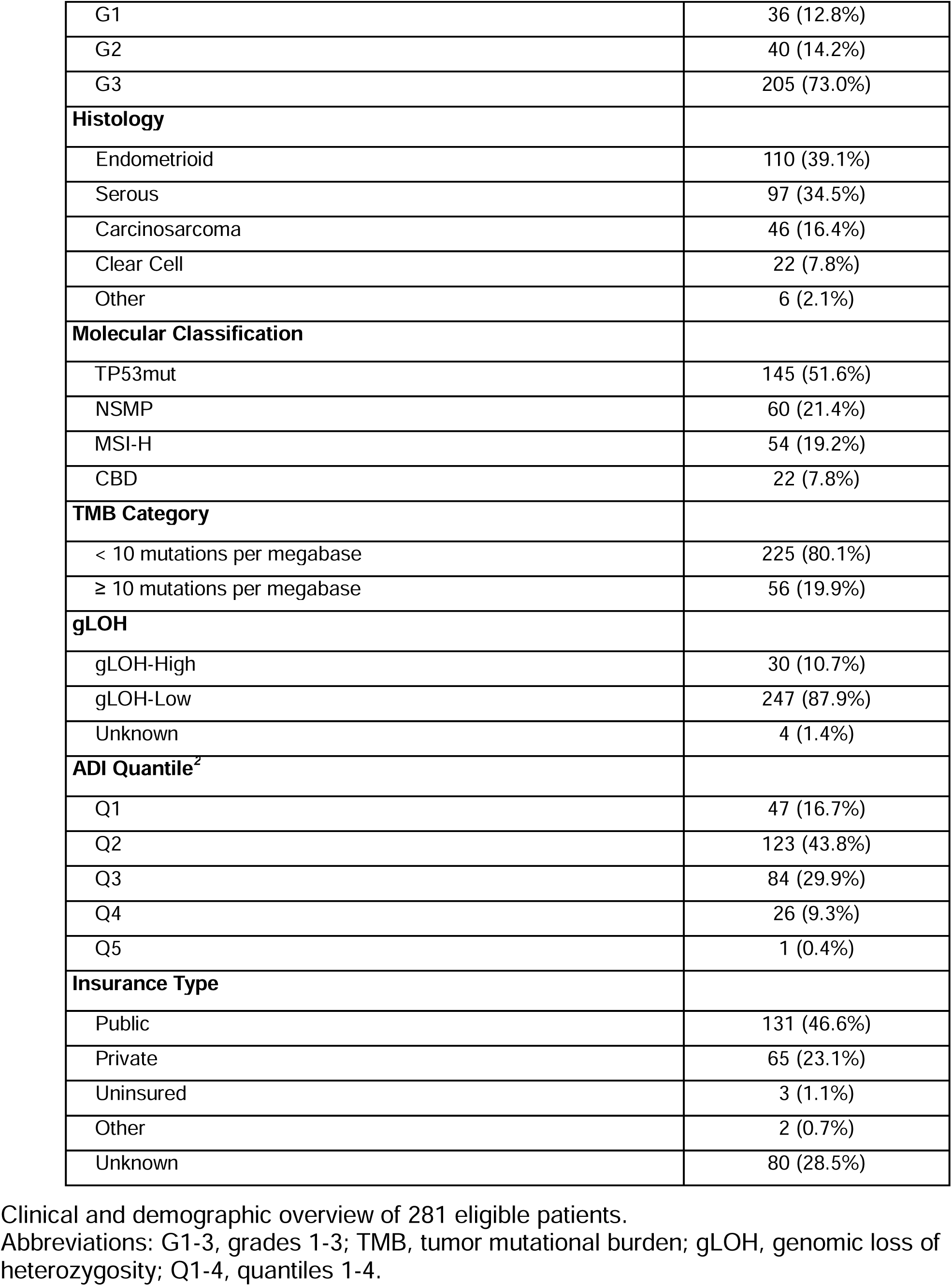
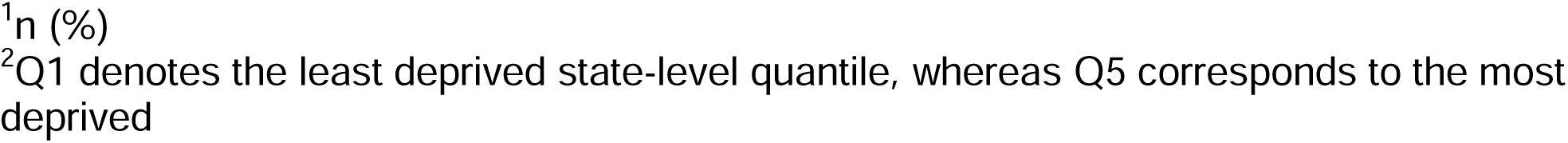
Patient Characteristics.

| **Characteristic** | **N = 281<sup>1</sup>** |
| --- | --- |
| **Age at Diagnosis** |  |
| <50 | 17 (6.0%) |
| 50-60 | 45 (16.0%) |
| 60-70 | 119 (42.3%) |
| 70-80 | 77 (27.4%) |
| 80+ | 23 (8.2%) |
| **Race** |  |
| White | 163 (58.0%) |
| Black or African American | 75 (26.7%) |
| Asian | 33 (11.7%) |
| Other | 10 (3.6%) |
| **Ethnicity** |  |
| Not Hispanic or Latino | 239 (85.1%) |
| Hispanic or Latino | 22 (7.8%) |
| Unknown | 20 (7.1%) |
| **Predominant Genetic Ancestry** |  |
| European | 148 (52.7%) |
| African | 76 (27.0%) |
| South Asian | 21 (7.5%) |
| Admixed American | 12 (4.3%) |
| East Asian | 11 (3.9%) |
| Unknown | 13 (4.6%) |
| **BMI Category** |  |
| Class 1 Obesity | 64 (22.8%) |
| Class 2 Obesity | 32 (11.4%) |
| Class 3 Obesity | 26 (9.3%) |
| Non-Obese | 141 (50.2%) |
| Unknown | 18 (6.4%) |
| **Stage** |  |
| I | 111 (39.5%) |
| II | 6 (2.1%) |
| III | 71 (25.3%) |
| IV | 93 (33.1%) |
| **Grade** |  |
| G1 | 36 (12.8%) |
| G2 | 40 (14.2%) |
| G3 | 205 (73.0%) |
| **Histology** |  |
| Endometrioid | 110 (39.1%) |
| Serous | 97 (34.5%) |
| Carcinosarcoma | 46 (16.4%) |
| Clear Cell | 22 (7.8%) |
| Other | 6 (2.1%) |
| **Molecular Classification** |  |
| TP53mut | 145 (51.6%) |
| NSMP | 60 (21.4%) |
| MSI-H | 54 (19.2%) |
| CBD | 22 (7.8%) |
| **TMB Category** |  |
| < 10 mutations per megabase | 225 (80.1%) |
| ≥ 10 mutations per megabase | 56 (19.9%) |
| **gLOH** |  |
| gLOH-High | 30 (10.7%) |
| gLOH-Low | 247 (87.9%) |
| Unknown | 4 (1.4%) |
| **ADI Quantile<sup>2</sup>** |  |
| Q1 | 47 (16.7%) |
| Q2 | 123 (43.8%) |
| Q3 | 84 (29.9%) |
| Q4 | 26 (9.3%) |
| Q5 | 1 (0.4%) |
| **Insurance Type** |  |
| Public | 131 (46.6%) |
| Private | 65 (23.1%) |
| Uninsured | 3 (1.1%) |
| Other | 2 (0.7%) |
| Unknown | 80 (28.5%) |

Clinical and demographic overview of 281 eligible patients. Abbreviations: G1-3, grades 1-3; TMB, tumor mutational burden; gLOH, genomic loss of heterozygosity; Q1-4, quantiles 1-4.
<sup>1</sup>n (%) <sup>2</sup>Q1 denotes the least deprived state-level quantile, whereas Q5 corresponds to the most deprived

We compared individuals of predominantly EUR or AFR genetic ancestry as there were limited sample sizes of other genetic ancestry groups in our cohort. In agreement with previous reports [7,26], AFR patients were more likely to have high-grade tumors as 89% of AFR patients had FIGO grade 3 tumors compared to 64% of EUR ancestry patients (p-value < 0.001) (**Table S1**). Among patients who could be assigned a molecular classification, AFR ancestry patients were more likely to be TP53mut compared to EUR ancestry patients (71% versus 51%, p-value = 0.015) (**Table S1**). The difference in subtype distribution largely stems from a higher prevalence of the NSMP molecular classification in EUR ancestry individuals, as there was not a statistically significant difference in microsatellite instability status between ancestry groups (**Table S1**). For patients with available BMI prior to surgery, AFR patients were more likely to be obese compared to EUR patients (62% versus 44%, p-value = 0.043) (**Table S1**). We did not observe significant differences in age or stage by genetic ancestry (**Table S1**). Genetic ancestry, grade, histology, and ADI quantiles were significantly different between White and Black or African American patients (**Table S2**).

### Histological Subtype and Molecular Classification Shape Ancestry-Associated Somatic Alterations

Key EC driver genes were frequently mutated in the cohort. Oncogenes such as *PIK3CA*, *KRAS*, *ERBB2*, and *CCNE1*, along with tumor suppressor genes such as *TP53*, *PTEN*, *ARID1A*, and *PIK3R1,* had the highest alteration frequencies (**Figure 2a**). *ERBB2*, *MYC*, and *CCNE1* were recurrently amplified in patients within this cohort, all of which are associated with worse outcomes [27] (**Figure 2a**). OncoKB [28,29] annotation of clinically actionable variants revealed significant differences in the actionability of variants between individuals of EUR and AFR genetic ancestry (**Figure S1**). AFR ancestry patients had a higher proportion of individuals with no actionable mutations, leading to fewer therapeutic options for these patients (**Figure S1**).

**Figure 2.**
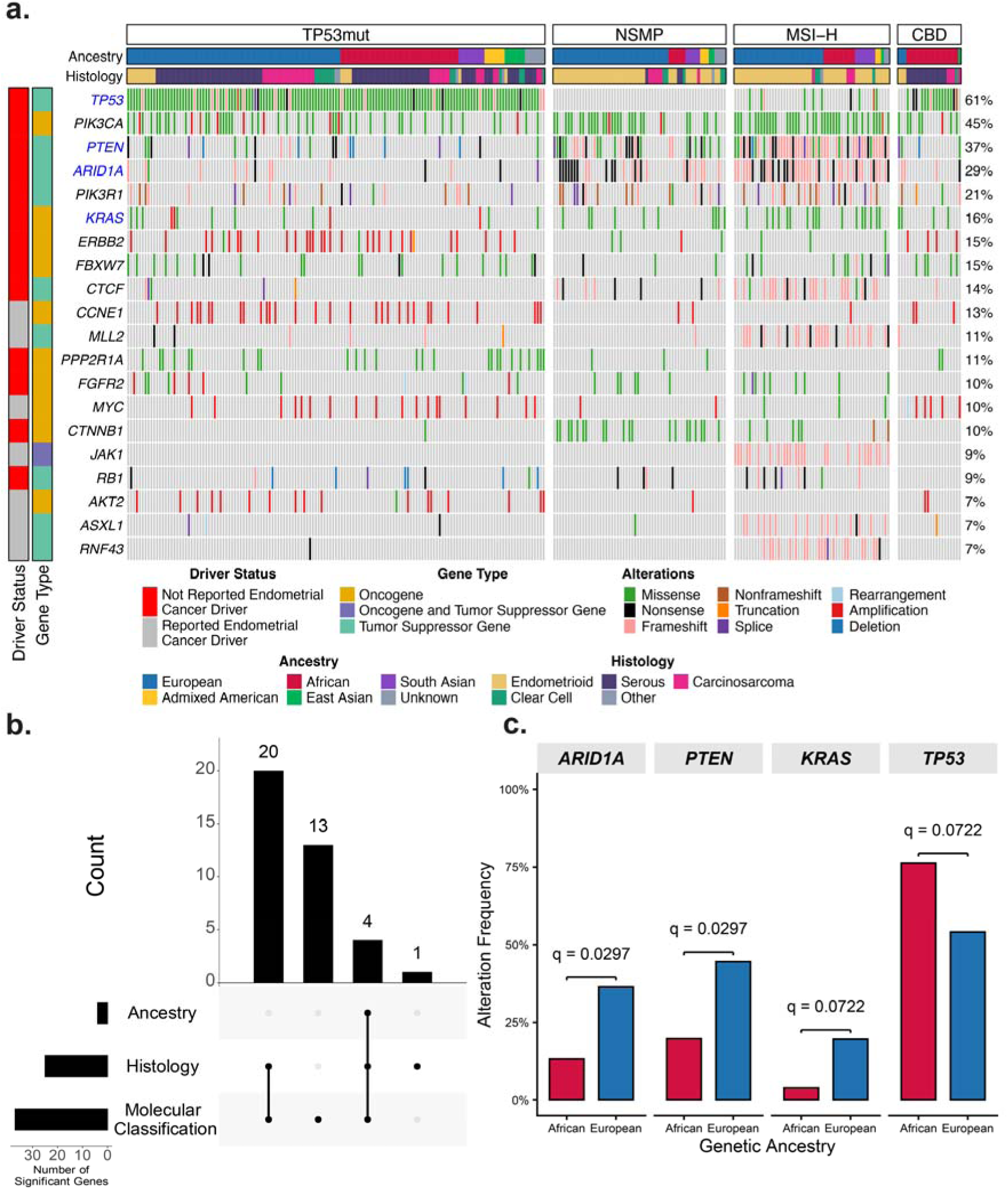
Association of Endometrial Cancer Somatic Alteration Landscape with Ancestry and Tumor Subtype: A) 213 genes had evidence of a pathogenic mutation in at least one patient. The top 20 most recurrently mutated genes are displayed. The driver status of genes is as reported from a comprehensive characterization of endometrial cancer driver mutations [39]. Gene types are displayed vertically, and patient characteristics are displayed horizontally. Gene names in blue were significantly different between genetic ancestry groups. B) Pairwise somatic alterations between groups defined by genetic ancestry, histology, or molecular classification revealed 38 distinct genes (q < 0.10, Fisher’s exact test). Distinct genes were frequently differentially mutated across multiple comparisons. The frequencies of overlapping comparisons are shown in the UpSet plot. C) *ARID1A*, *PTEN*, *KRAS*, and *TP53* had significantly different alteration frequencies between genetic ancestry groups.

Pairwise comparisons between groups defined by molecular classifications and histological subtypes revealed 37 genes with significantly different alteration frequencies between molecular classifications and 25 genes between histological subtypes (**Figure 2b**, **Tables S3 and S4**). 4 genes, *TP53*, *ARID1A*, *PTEN*, and *KRAS* were significantly different between individuals of EUR and AFR genetic ancestry (q-value < 0.10) (**Figure 2c, Table S5**). Each gene was also significantly different between molecular classifications and histological subtypes (**Figure 2b**). However, this is likely driven by differences in the distribution of molecular classifications and histological subtypes between ancestry groups (**Table 1**), since comparison of somatic alterations by ancestry stratified by molecular classification yielded no significant results. Somatic alterations in genes frequently mutated in cancer may be more strongly associated with histological subtype and molecular classification rather than ancestry.

### State Level Area Deprivation is Not Associated with Worse Clinical Outcomes

To facilitate group comparisons, we collapsed New York state ADI quantiles into four groups since only a minority of patients come from the most deprived region, quantile 5 (Q5). Patients were primarily from less deprived socioeconomic regions as 60.5% of the patients resided in Q1 and Q2 (**Table 1**). Race was significantly associated with ADI quantiles, while genetic ancestry groups were not correlated with area deprivation (**Table S6**). The least deprived ADI quantiles had more White and Asian patients, while the more deprived quantiles had more Black or African American and Other patients (**Table S6**, **Figure 3**). All other factors under investigation, including tumor characteristics (stage, grade, molecular classification, histology, microsatellite instability, tumor mutational burden) or demographic variables were not associated with ADI quantiles (**Table S6**).

**Figure 3.**
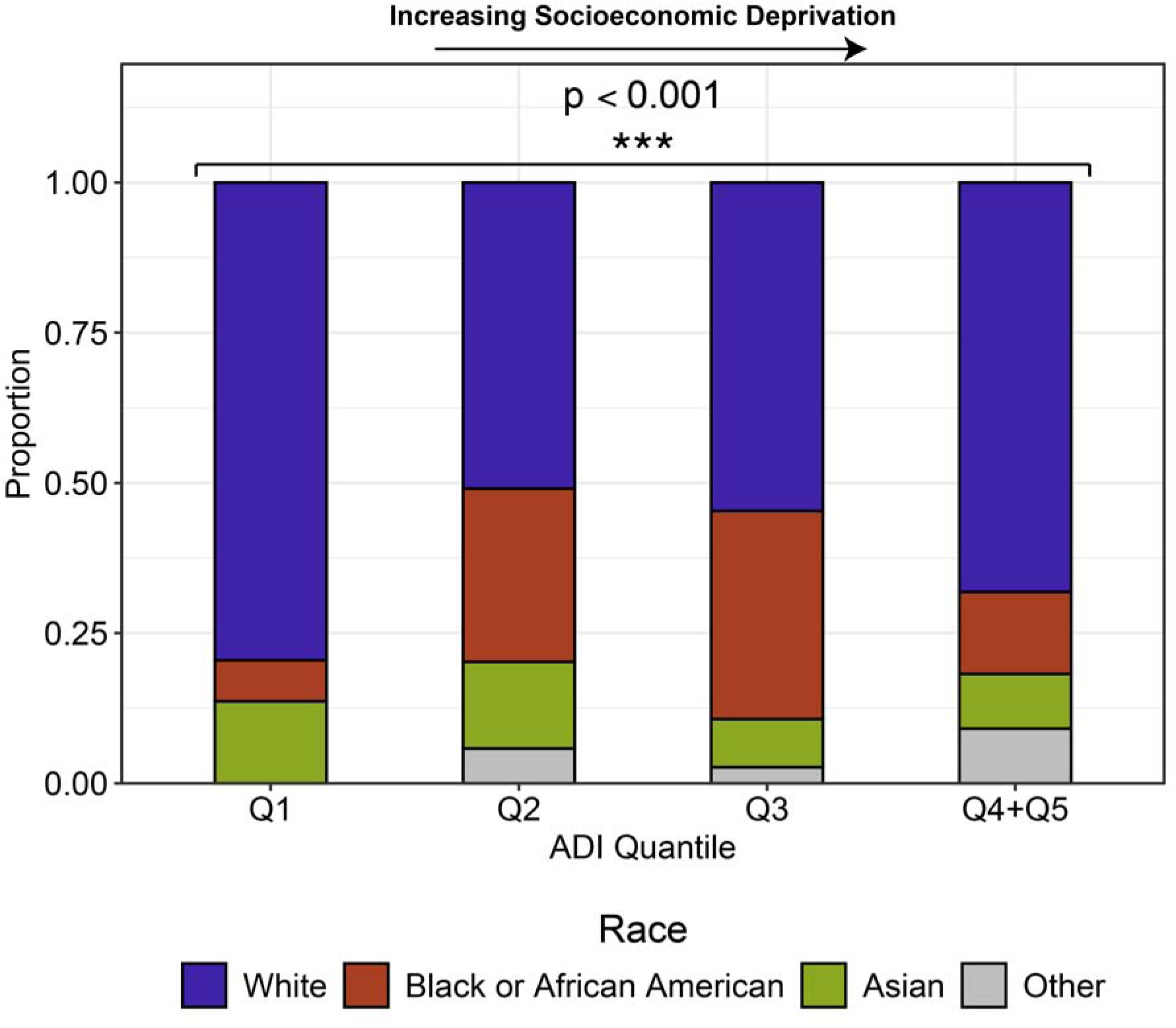
Distribution of Racial Groups Across Area Deprivation Quantiles: The proportion of each self-identified race across ADI state quantiles is displayed. Q1 represents the most advantaged socioeconomic region while Q4+Q5 represents the most deprived.

In univariate analyses, both race and genetic ancestry were significantly associated with PFS (Race p-value = 0.03; Ancestry p-value = 0.02) and OS (Race p-value = 0.01; Ancestry p-value = 0.01) (**Figure 4, Figure S2**). Patients identifying as Black or African American, and patients with predominantly African ancestry had worse PFS and OS compared to patients identifying as White or predominantly European ancestry (**Figure 4**). Patients identifying as Asian and patients with East or South Asian ancestry did not have worse PFS and OS compared to White or European ancestry patients (**Figure 4, Figure S2**). Overall, ADI quantiles were not significantly associated with PFS or OS (PFS p-value = 0.09, OS p-value = 0.68) (**Figure S2**).

**Figure 4.**
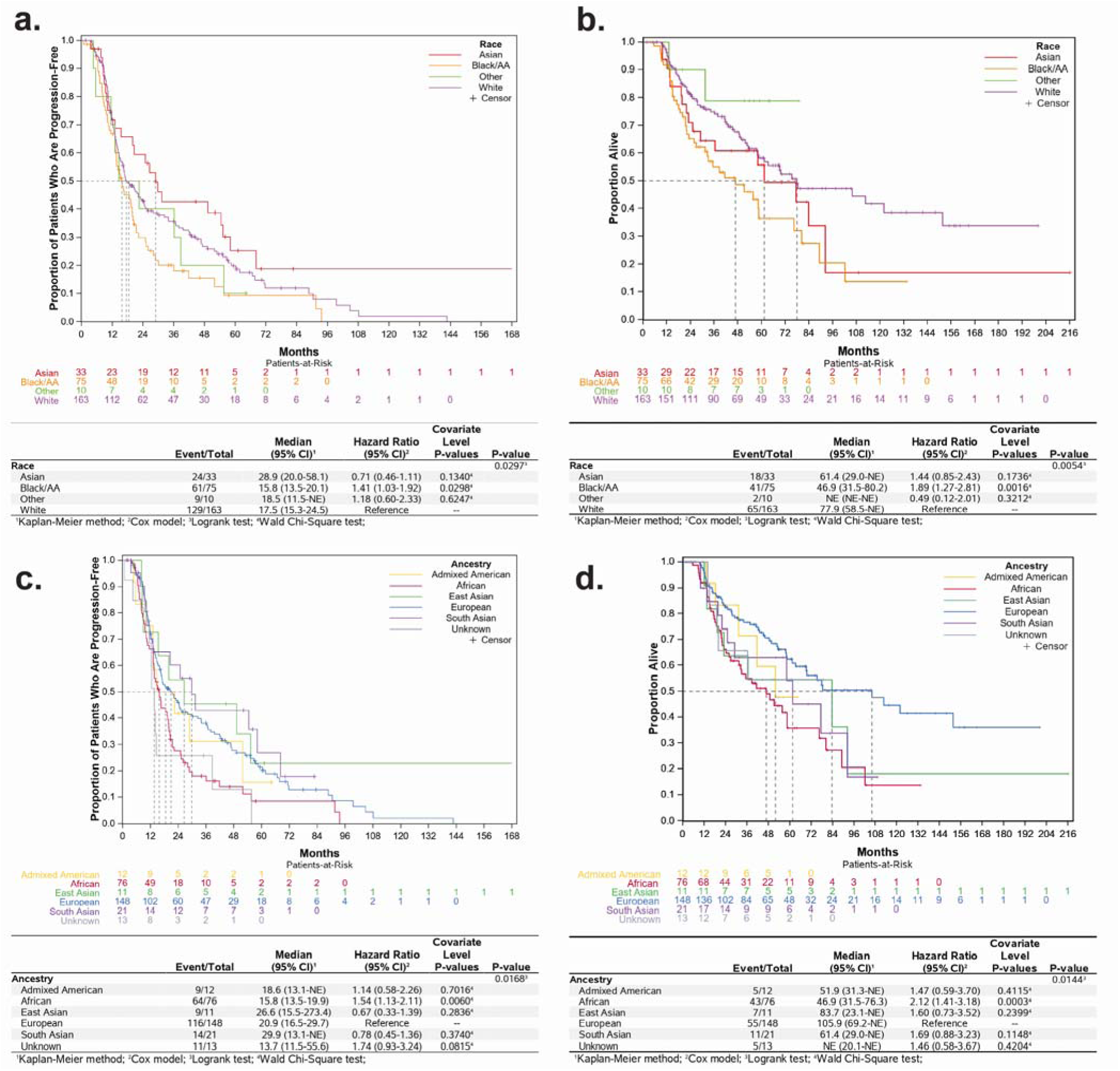
Kaplan-Meier Survival Curves for Progression-Free and Overall Survival by Race (A-B) and Ancestry (C-D): The Kaplan-Meier method was used to estimate survival distributions, and differences between groups were assessed using the log-rank test.

### African Genetic Ancestry is Associated with Poorer Clinical Outcomes

A Cox proportional hazards (CPH) model was constructed by combining socioeconomic, clinical, and molecular factors for the 241 patients with the necessary data (**Figure 1**). MSI-H and NSMP molecular classifications had better PFS (MSI-H HR 0.36, p-value = 0.0002; NSMP HR 0.6, p-value = 0.0377) and OS (MSI-H HR 0.25, p-value = 0.001; NSMP HR 0.45, p-value = 0.0159) compared to TP53mut molecular classification (**Figure 5, Figure S3**). Stages III and IV disease were significantly associated with poorer OS (HR 2.04, p-value = 0.001) (**Figure S3**). ADI quantiles did not have significantly different PFS or OS (**Figure 5**, **Figure S3**). Despite controlling for a spectrum of factors that affect survival, AFR patients had statistically significant poorer PFS and OS compared to EUR ancestry patients (PFS HR 1.91, p-value = 0.0013; OS HR 1.72, p-value = 0.0322) (**Figure 5**). In CPH models that substituted ancestry for race, there were similar associations between cohort characteristics and survival, with the exception of the OS model (**Table S9 and S10**). In this model, Black or African American patients did not have significantly different OS compared to White patients (HR 1.54, p-value = 0.0876) (**Table S9 and S10**).

**Figure 5.**
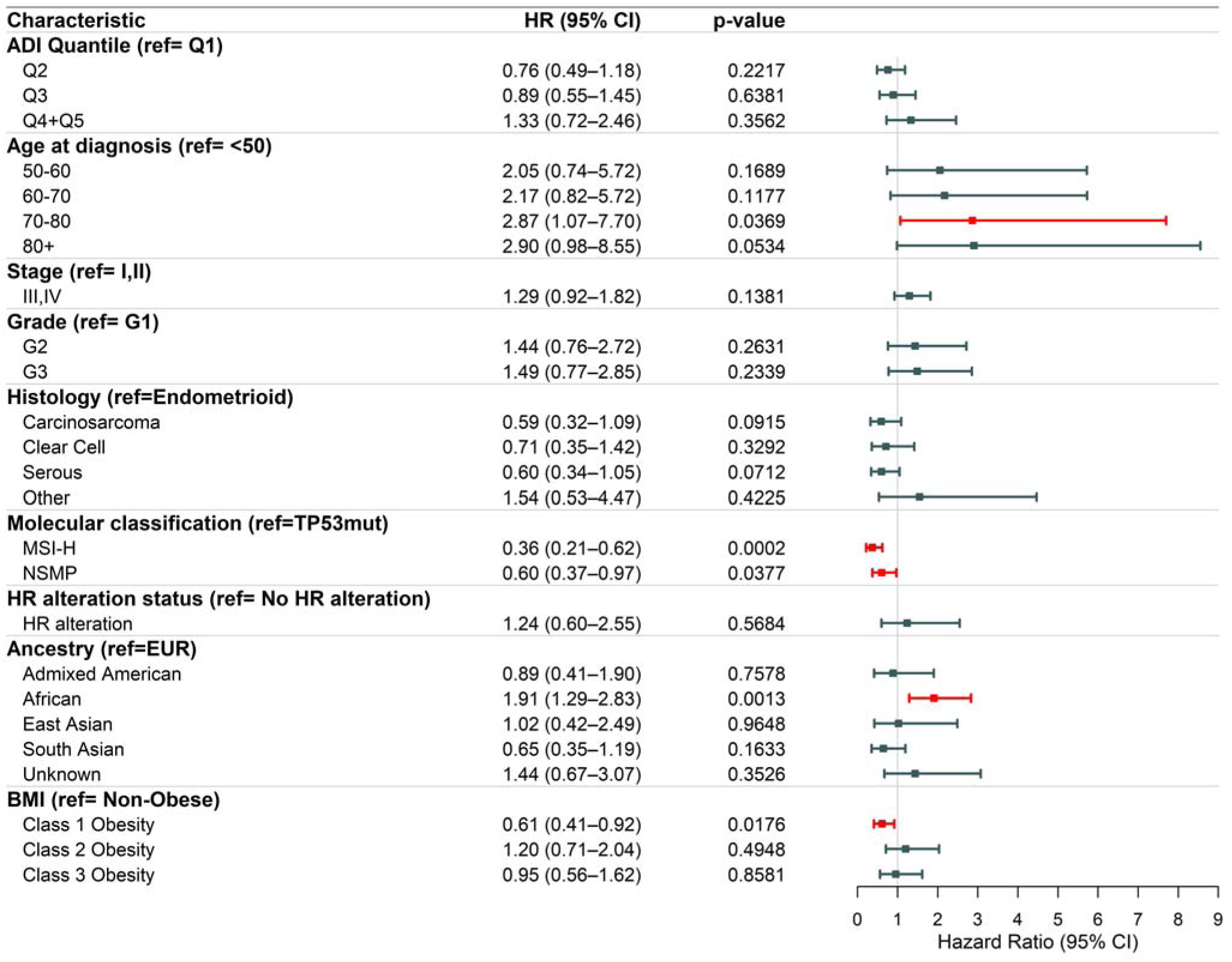
Multivariable Cox Proportional Hazards Model for Progression-Free Survival: A Cox proportional hazards regression model incorporated covariates that were either clinically relevant or statistically significant in univariable analyses. Significant differences are shown in red.

## Discussion

This study analyzed a diverse group of women with predominantly high-grade EC. Compared to women of EUR ancestry, AFR ancestry women had more aggressive disease, characterized by higher grade, non-endometrioid tumors, and clinical features associated with poorer prognosis. Somatic alterations differed significantly between genetic ancestry groups due to differences in the distribution of histological subtypes and molecular classifications. Groups defined by race were differentially distributed across state-level ADI quantiles. Despite this association, these quantiles were not associated with survival outcomes, whereas race and ancestry groupings showed significantly different PFS and OS outcomes. Including each of these factors into multivariable survival models revealed persistent disparities in outcomes between AFR and EUR ancestry women. These results suggest that features other than access to care, state-level ADI, histological subtype, and molecular classification may account for the extent of this disparity.

Our results recapitulated the observed EC trends, in which molecular features associated with poor prognosis are more frequent in AFR women compared to women of EUR [7,26]. In our cohort, somatic alterations in *TP53*, a prognostic marker of aggressive disease, were associated with AFR ancestry. Additionally, somatic alterations associated with better prognosis in high-grade EC such as *PTEN* and *ARID1A* were associated with EUR ancestry [30,31]. Similar to the associations between pathogenic germline variants and molecular classifications that have been previously reported [32], we found a correlation between molecular classifications and somatic alterations. These data illustrate that molecular classification likely groups individuals with similar somatic and germline pathogenic variation in genes frequently altered in EC. As a result, including molecular classifications in our survival model allows us to account for variation in these commonly studied genes. Additional investigation into germline and somatic variation beyond common oncogenic drivers may provide a better understanding of the incidence trends of EC subtypes and responses to treatment that contribute to disparities by ancestry. Ancestry-related EC biological differences have been reported in DNA methylation and the tumor immune microenvironment [33–36]. As each of these biological mechanisms can be influenced by environmental factors, it is important for future endometrial studies to include the full spectrum of potential drivers of mortality and model their interactions to better understand EC disparities.

In this study, we found that although racial groups were differentially distributed across New York state ADI quantiles, these quantiles did not affect differences in PFS or OS. This finding is broadly in line with another New York City based study that reported that ADI has no significant association with quality of care or overall survival within the New York metropolitan area [37]. These findings may reflect the frequency of mixed-status neighborhoods in New York City, and the summarized nature of ADI. Another possible reason is that in this dense urban area, there are fewer geographic barriers to receiving care, even for patients with limited resources. Since our cohort is primarily from the New York metropolitan area and received care at different Northwell clinics, the selected patients likely had similar access to care across ADI quantiles. Our results did not indicate a relationship between ADI and tumor biology. While there may be an association between tumor biology and ADI on a national level, our results suggest that there are likely other factors that drive an increased frequency of non-endometrioid aggressive tumors in Black women.

On a national level, it has been demonstrated that the protective effect of socioeconomic advantage in EC survival is differential between racial groups [38]. White women in more advantaged socioeconomic regions have better overall survival compared to White women in more deprived socioeconomic regions, while this relationship does not exist for Black women. This creates a larger difference in survival between Black and White women in more advantaged socioeconomic regions, as one group benefits from affluence while the other does not [38]. The majority of patients in this study belong to the top two quantiles of New York State. Thus, our study is composed of women for whom differences in survival between Black and White patients within socioeconomic regions are the highest. Our results indicate that characteristics of tumor biology such as molecular classification also do not explain this difference.

Multiple limitations were identified, including the retrospective nature of this cohort, and patients included in this study were limited to those who underwent testing using specific CGP assays; thus, sampling bias limits the study’s representativeness (Figure 1). Although tumor genomic testing is now the standard of care for most high-grade tumors at diagnosis, previously patients received sequencing only if they had recurrent, high-grade disease. Since recurrent disease is more common among Black women, we believe that our findings still reflect the subset of tumors where disparities are the greatest. Additionally, race was extracted from patients’ electronic medical records, and may have been self-identified by patients, or recorded by a provider. Though genetic ancestry is continuous, we use patients’ predominant genetic ancestry as a categorical variable due to limitations in our genetic ancestry inference methods. We acknowledge that though this does not fully depict the complexities of genetic ancestry, we still seek to estimate the populations that are disproportionately burdened by endometrial cancer risk, mortality, and their associated characteristics. While ideally, we would simultaneously model both race and ancestry in evaluating survival, the variables were highly correlated. This correlation makes it impossible for the CPH model to disentangle their independent effects. This would lead to unstable hazard ratio estimates, inflated standard errors, and an uninterpretable model. For these reasons we could not jointly model race and ancestry.

Despite these limitations, we were able to combine a wide range of previously siloed factors to model EC survival in a large, diverse cohort of predominantly high-grade EC patients. Given evidence of differences in mortality being highest in more advantaged socioeconomic settings, our study captures a subset of women where differences in EC survival are the most extreme.

Our findings indicate that EC disparities result from a complex interplay of factors. Even after adjusting for grade, histology, advanced stage, and ADI, African ancestry remained independently associated with poorer PFS and OS. While EC disparities are certainly driven by unequal access to care, socioeconomic deprivation, and the frequency of poor prognostic tumor characteristics in Black women, there are still components of this disparity that are not explained by these differences. This points towards the need for further examination of the systemic and individual barriers Black women experience that may affect their health outcomes. Additionally, a better understanding of the interplay of ancestry and molecular characteristics will lead to the development of targeted therapeutic approaches that will reduce persisting disparities.

## Contributions

**Devin Gee**: Conceptualization, Data Curation, Formal Analysis, Investigation, Methodology, Project Administration, Software, Visualization, Writing – Original Draft Preparation, Writing – Review and Editing. **Alisha Daroch**: Data Curation, Investigation, Writing – Original Draft Preparation, Writing – Review and Editing. **Meredith Akerman**: Formal Analysis, Investigation, Methodology, Software, Visualization, Writing – Original Draft Preparation, Writing – Review and Editing. **Natalie Danziger**: Data Curation, Investigation, Methodology, Resources, Writing – Original Draft Preparation, Writing – Review and Editing. **Leslie Panella**: Data Curation, Investigation, Methodology, Writing – Review and Editing. **Megan Gorman**: Data Curation, Investigation, Methodology, Writing – Review and Editing. **Madeline Bright**: Data Curation, Investigation, Methodology, Writing – Review and Editing. **Douglas I. Lin**: Investigation, Methodology, Resources, Writing – Review and Editing. **Nyasha Chambwe**: Conceptualization, Project Administration, Resources, Supervision, Writing – Review and Editing. **Marina Frimer**: Conceptualization, Data Curation, Investigation, Project Administration, Resources, Supervision, Writing – Original Draft, Writing – Review and Editing.

## Data Availability

All data produced in the present work are contained in the manuscript. If needed, additional details may also be obtained by contacting the corresponding authors.

