## Supplemental Figures and Tables for "Endometrial cancer survival disparities in women of African ancestry persist beyond clinical, molecular, and socioeconomic determinants"

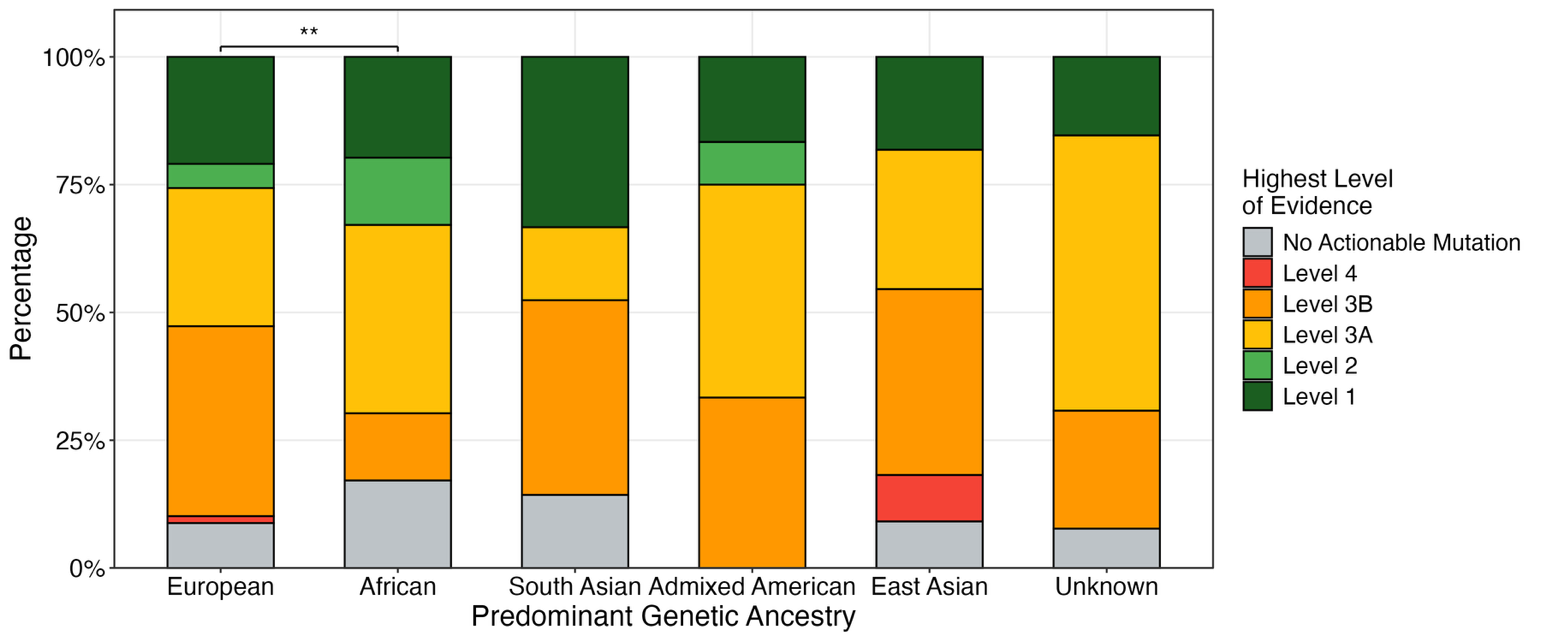

**Figure S1. OncoKB [28,29] Annotation of Clinical Actionability by Genetic Ancestry:** The clinical actionability of single nucleotide, copy number, structural, and atypical variants were annotated using the OncoKB database. Level 1 represents a biomarker for standard of care treatment (high actionability), while Level 4 represents a hypothetical biomarker (low actionability).

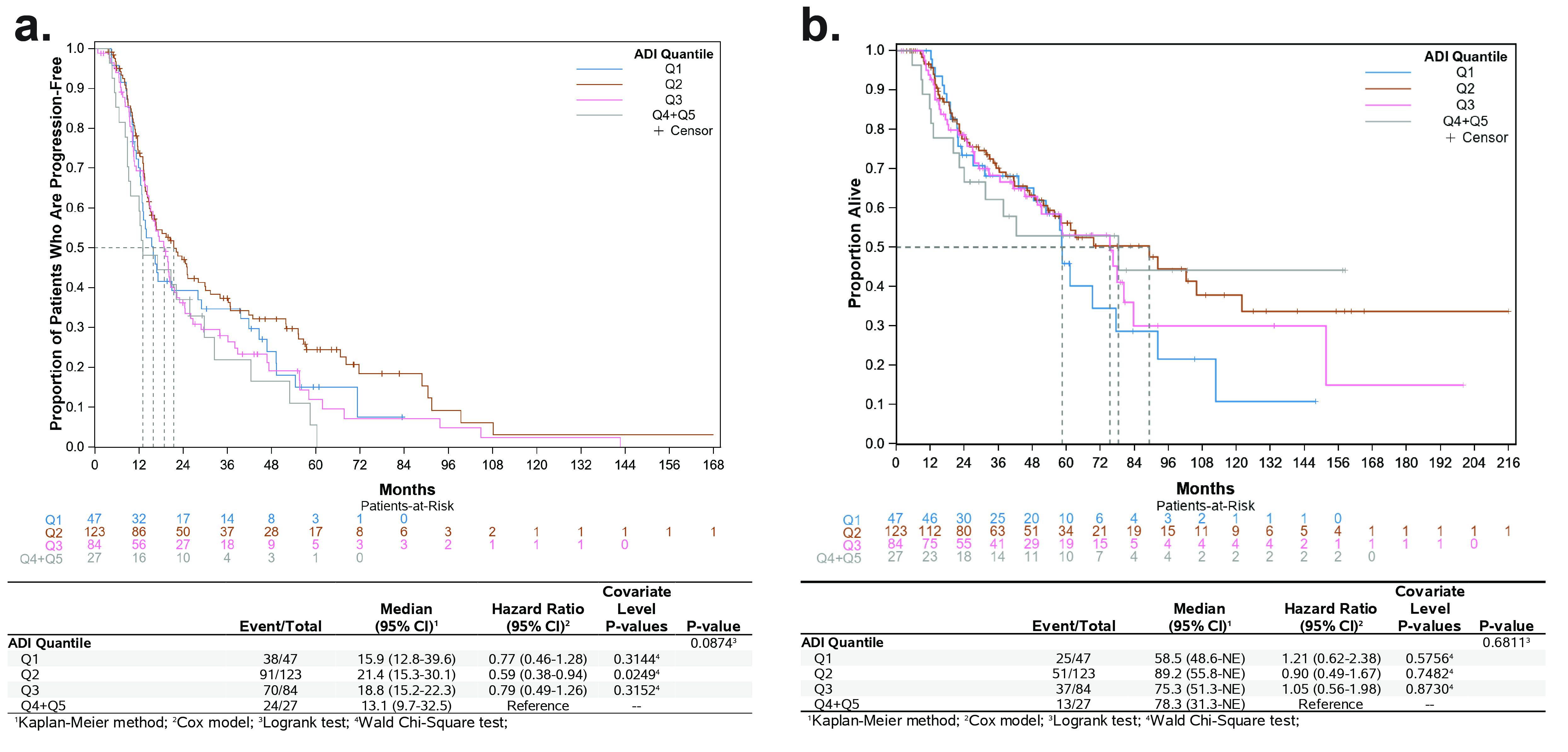

**Figure S2. Kaplan-Meier Plot for Progression-Free and Overall Survival by ADI:** The Kaplan-Meier method was used to estimate survival distributions, and differences between groups were assessed using the log-rank test.

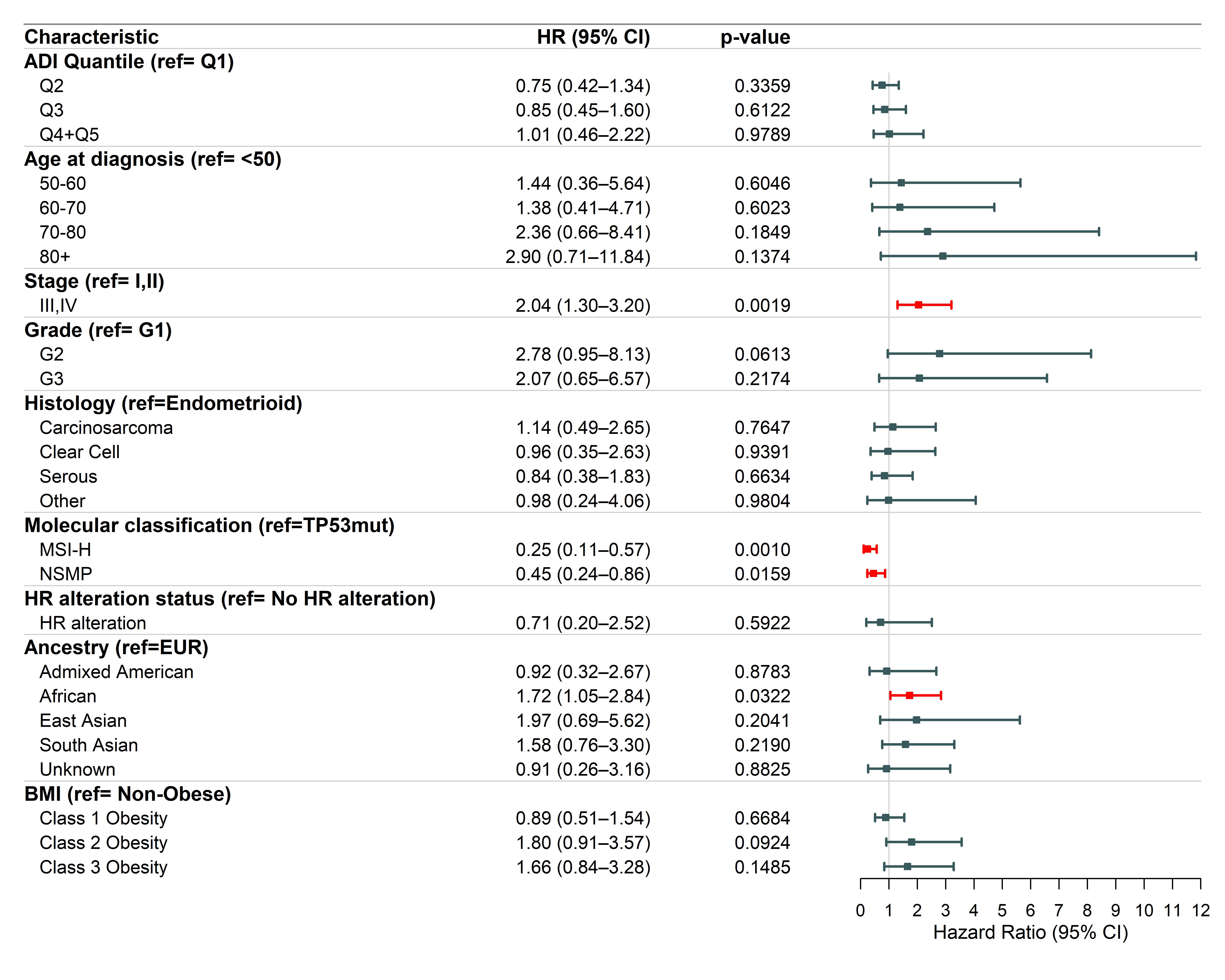

**Figure S3. Multivariable Cox Proportional Hazards Model for Overall Survival:** A Cox proportional hazards regression model incorporated covariates that were either clinically relevant or statistically significant in univariable analyses. Significant differences are shown in red.

**Table S1. Comparison of Cohort Characteristics by Genetic Ancestry**

| **Characteristic** | **European** N = 148*^1^* | **African** N = 76*^1^* | **p-value***^2^* |
| --- | --- | --- | --- |
| **Age at Diagnosis** |  |  | 0.10 |
| <50 | 11 (7.4%) | 0 (0%) |  |
| 50-60 | 24 (16%) | 11 (14%) |  |
| 60-70 | 55 (37%) | 35 (46%) |  |
| 70-80 | 44 (30%) | 21 (28%) |  |
| 80+ | 14 (9.5%) | 9 (12%) |  |
| **Race** |  |  | <0.001 |
| White | 146 (99%) | 4 (5.3%) |  |
| Black or African American | 1 (0.7%) | 72 (95%) |  |
| Asian | 0 (0%) | 0 (0%) |  |
| Other | 1 (0.7%) | 0 (0%) |  |
| **Ethnicity** |  |  | 0.9 |
| Not Hispanic or Latino | 133 (90%) | 67 (88%) |  |
| Hispanic or Latino | 3 (2.0%) | 2 (2.6%) |  |
| Unknown | 12 (8.1%) | 7 (9.2%) |  |
| **BMI Category** |  |  | 0.043 |
| Non-Obese | 76 (56%) | 29 (38%) |  |
| Class 1 Obesity | 29 (21%) | 29 (38%) |  |
| Class 2 Obesity | 17 (13%) | 10 (13%) |  |
| Class 3 Obesity | 14 (10%) | 8 (11%) |  |
| Unknown | 12 | 0 |  |
| **Stage** |  |  | 0.5 |
| I | 58 (39%) | 30 (39%) |  |
| II | 4 (2.7%) | 2 (2.6%) |  |
| III | 43 (29%) | 16 (21%) |  |
| IV | 43 (29%) | 28 (37%) |  |
| **Grade** |  |  | <0.001 |
| G1 | 26 (18%) | 4 (5.3%) |  |
| G2 | 28 (19%) | 4 (5.3%) |  |
| G3 | 94 (64%) | 68 (89%) |  |
| **Molecular Classification** |  |  | 0.015 |
| TP53mut | 74 (51%) | 41 (71%) |  |
| NSMP | 40 (28%) | 6 (10%) |  |
| MSI-H | 31 (21%) | 11 (19%) |  |
| Unknown | 3 | 18 |  |
| **Histology** |  |  | <0.001 |
| Endometrioid | 72 (49%) | 15 (20%) |  |
| Serous | 38 (26%) | 43 (57%) |  |
| Carcinosarcoma | 24 (16%) | 13 (17%) |  |
| Clear Cell | 11 (7.4%) | 4 (5.3%) |  |
| Other | 3 (2.0%) | 1 (1.3%) |  |
| **Microsatellite Instability** |  |  | 0.7 |
| MSI-H | 31 (21%) | 11 (19%) |  |
| MSS | 114 (79%) | 47 (81%) |  |
| Unknown | 3 | 18 |  |
| **TMB Category** |  |  | 0.6 |
| < 10 mutations per megabase | 118 (80%) | 63 (83%) |  |
| ≥ 10 mutations per megabase | 30 (20%) | 13 (17%) |  |
| **gLOH** |  |  | 0.5 |
| gLOH-High | 13 (8.8%) | 9 (12%) |  |
| gLOH-Low | 132 (89%) | 67 (88%) |  |
| Unknown | 3 (2.0%) | 0 (0%) |  |

Comparison of demographic and clinical characteristics between 148 patients of predominantly European genetic ancestry and 76 patients of predominantly African genetic ancestry.

Abbreviations: G1-3, grades 1-3; TMB, tumor mutational burden; gLOH, genomic loss of heterozygosity.

^1^n (%)

^2^Fisher's Exact Test for Count Data with simulated p-value (based on 2000 replicates); Pearson's Chi-squared test

**Table S2. Comparison of Cohort Characteristics by Race**

| **Characteristic** | **White** N = 163*^1^* | **Black or African American** N = 75*^1^* | **p-value***^2^* |
| --- | --- | --- | --- |
| **Age at Diagnosis** |  |  | 0.058 |
| <50 | 13 (8.0%) | 0 (0%) |  |
| 50-60 | 27 (17%) | 10 (13%) |  |
| 60-70 | 61 (37%) | 37 (49%) |  |
| 70-80 | 47 (29%) | 20 (27%) |  |
| 80+ | 15 (9.2%) | 8 (11%) |  |
| **Predominant Genetic Ancestry** |  |  | <0.001 |
| European | 146 (90%) | 1 (1.3%) |  |
| African | 4 (2.5%) | 72 (96%) |  |
| South Asian | 0 (0%) | 0 (0%) |  |
| Admixed American | 6 (3.7%) | 0 (0%) |  |
| East Asian | 0 (0%) | 0 (0%) |  |
| Unknown | 7 (4.3%) | 2 (2.7%) |  |
| **Ethnicity** |  |  | 0.2 |
| Not Hispanic or Latino | 140 (86%) | 67 (89%) |  |
| Hispanic or Latino | 11 (6.7%) | 1 (1.3%) |  |
| Unknown | 12 (7.4%) | 7 (9.3%) |  |
| **BMI Category** |  |  | 0.055 |
| Non-Obese | 84 (56%) | 29 (39%) |  |
| Class 1 Obesity | 31 (21%) | 27 (36%) |  |
| Class 2 Obesity | 19 (13%) | 10 (14%) |  |
| Class 3 Obesity | 15 (10%) | 8 (11%) |  |
| Unknown | 14 | 1 |  |
| **Stage** |  |  | 0.5 |
| I | 65 (40%) | 28 (37%) |  |
| II | 4 (2.5%) | 2 (2.7%) |  |
| III | 46 (28%) | 16 (21%) |  |
| IV | 48 (29%) | 29 (39%) |  |
| **Grade** |  |  | 0.001 |
| G1 | 27 (17%) | 4 (5.3%) |  |
| G2 | 30 (18%) | 5 (6.7%) |  |
| G3 | 106 (65%) | 66 (88%) |  |
| **Molecular Classification** |  |  | 0.051 |
| TP53mut | 85 (53%) | 40 (70%) |  |
| NSMP | 42 (26%) | 7 (12%) |  |
| MSI-H | 33 (21%) | 10 (18%) |  |
| Unknown | 3 | 18 |  |
| **Histology** |  |  | <0.001 |
| Endometrioid | 77 (47%) | 14 (19%) |  |
| Serous | 42 (26%) | 43 (57%) |  |
| Carcinosarcoma | 26 (16%) | 14 (19%) |  |
| Clear Cell | 14 (8.6%) | 3 (4.0%) |  |
| Other | 4 (2.5%) | 1 (1.3%) |  |
| **TMB Category** |  |  | 0.5 |
| < 10 mutations per megabase | 131 (80%) | 63 (84%) |  |
| ≥ 10 mutations per megabase | 32 (20%) | 12 (16%) |  |
| **gLOH** |  |  | 0.4 |
| gLOH-High | 16 (9.8%) | 9 (12%) |  |
| gLOH-Low | 143 (88%) | 66 (88%) |  |
| Unknown | 4 (2.5%) | 0 (0%) |  |
| **ADI Quantile** |  |  | <0.001 |
| Q1 | 37 (23%) | 3 (4.0%) |  |
| Q2 | 63 (39%) | 37 (49%) |  |
| Q3 | 44 (27%) | 31 (41%) |  |
| Q4+Q5 | 19 (12%) | 4 (5.3%) |  |
| **Insurance Type** |  |  | 0.5 |
| Public | 82 (50%) | 30 (40%) |  |
| Private | 33 (20%) | 21 (28%) |  |
| Uninsured | 2 (1.2%) | 1 (1.3%) |  |
| Other | 1 (0.6%) | 0 (0%) |  |
| Unknown | 45 (28%) | 23 (31%) |  |

Comparison of demographic and clinical characteristics between 163 White patients and 75 Black or African American patients.

Abbreviations: G1-3, grades 1-3; TMB, tumor mutational burden; gLOH, genomic loss of heterozygosity.

^1^n (%)

^2^Fisher's Exact Test for Count Data with simulated p-value (based on 2000 replicates); Pearson's Chi-squared test

**Table S3. Pairwise Comparisons of Significant Genes Between Molecular Classifications**

Pairwise comparisons by molecular classification

| **gene** | **comparison** | **odds_ratio** | **p** | **q** |
| --- | --- | --- | --- | --- |
| *TP53* | TP53mut vs NSMP | 35211.00 | 2.46e-53 | 5.25e-51 |
| *TP53* | TP53mut vs MSI-H | 1233.29 | 7.73e-35 | 1.65e-32 |
| *PTEN* | TP53mut vs MSI-H | 0.02 | 6.93e-22 | 7.38e-20 |
| *ARID1A* | TP53mut vs MSI-H | 0.04 | 1.25e-17 | 8.88e-16 |
| *JAK1* | TP53mut vs MSI-H | 0.00 | 4.25e-17 | 2.27e-15 |
| *RNF43* | TP53mut vs MSI-H | 0.01 | 3.81e-12 | 1.62e-10 |
| *TP53* | NSMP vs CBD | 0.00 | 1.79e-12 | 3.81e-10 |
| *CTCF* | TP53mut vs MSI-H | 0.04 | 1.19e-11 | 4.21e-10 |
| *MLL2* | TP53mut vs MSI-H | 0.05 | 4.93e-11 | 1.50e-09 |
| *CTNNB1* | TP53mut vs NSMP | 0.01 | 2.58e-11 | 2.75e-09 |
| *JAK1* | NSMP vs MSI-H | 0.01 | 1.69e-10 | 3.61e-08 |
| *PTEN* | MSI-H vs CBD | 36.00 | 5.15e-09 | 1.10e-06 |
| *MLL2* | NSMP vs MSI-H | 0.02 | 1.39e-08 | 1.48e-06 |
| *ASXL1* | TP53mut vs MSI-H | 0.05 | 6.82e-08 | 1.82e-06 |
| *RNF43* | NSMP vs MSI-H | 0.01 | 3.34e-08 | 2.37e-06 |
| *QKI* | TP53mut vs MSI-H | 0.01 | 1.12e-06 | 2.66e-05 |
| *ARID1A* | TP53mut vs NSMP | 0.15 | 6.77e-07 | 4.81e-05 |
| *BCORL1* | TP53mut vs MSI-H | 0.02 | 4.74e-06 | 8.41e-05 |
| *MSH2* | TP53mut vs MSI-H | 0.02 | 4.74e-06 | 8.41e-05 |
| *PTCH1* | TP53mut vs MSI-H | 0.02 | 4.74e-06 | 8.41e-05 |
| *PTEN* | TP53mut vs NSMP | 0.20 | 2.93e-06 | 0.000156 |
| *ATM* | TP53mut vs MSI-H | 0.03 | 9.73e-06 | 0.000159 |
| *PTEN* | NSMP vs MSI-H | 0.12 | 3.08e-06 | 0.000164 |
| *BCOR* | TP53mut vs MSI-H | 0.07 | 1.35e-05 | 0.000205 |
| *HNF1A* | TP53mut vs MSI-H | 0.02 | 1.97e-05 | 0.000262 |
| *MSH6* | TP53mut vs MSI-H | 0.02 | 1.97e-05 | 0.000262 |
| *TP53* | TP53mut vs CBD | 114.64 | 2.71e-06 | 0.000578 |
| *PIK3CA* | TP53mut vs MSI-H | 0.25 | 4.68e-05 | 0.000586 |
| *NFE2L2* | TP53mut vs MSI-H | 0.02 | 8.04e-05 | 0.000951 |
| *ASXL1* | NSMP vs MSI-H | 0.04 | 2.29e-05 | 0.000974 |
| *TP53* | MSI-H vs CBD | 0.09 | 1.26e-05 | 0.001340 |
| *ARID1A* | MSI-H vs CBD | 11.70 | 2.40e-05 | 0.001433 |
| *JAK1* | MSI-H vs CBD | 38.90 | 2.69e-05 | 0.001433 |
| *CREBBP* | TP53mut vs MSI-H | 0.04 | 0.000139 | 0.001560 |
| *PPP2R1A* | TP53mut vs MSI-H | 24.17 | 0.000213 | 0.002272 |
| *MAP3K1* | TP53mut vs MSI-H | 0.03 | 0.000323 | 0.003277 |
| *KRAS* | TP53mut vs MSI-H | 0.23 | 0.000363 | 0.003513 |
| *CTNNB1* | TP53mut vs MSI-H | 0.05 | 0.000505 | 0.004678 |
| *CCND1* | TP53mut vs MSI-H | 0.08 | 0.000547 | 0.004851 |
| *CCNE1* | TP53mut vs MSI-H | 13.83 | 0.000658 | 0.005605 |
| *CTCF* | NSMP vs MSI-H | 0.19 | 0.000318 | 0.009362 |
| *TP53* | NSMP vs MSI-H | 0.04 | 0.000352 | 0.009362 |
| *QKI* | NSMP vs MSI-H | 0.04 | 0.000352 | 0.009362 |
| *SOX9* | TP53mut vs MSI-H | 0.03 | 0.001279 | 0.010480 |
| *SETD2* | TP53mut vs MSI-H | 0.06 | 0.001780 | 0.013588 |
| *EP300* | TP53mut vs MSI-H | 0.09 | 0.001786 | 0.013588 |
| *BCORL1* | NSMP vs MSI-H | 0.04 | 0.000820 | 0.015887 |
| *MSH2* | NSMP vs MSI-H | 0.04 | 0.000820 | 0.015887 |
| *PTCH1* | NSMP vs MSI-H | 0.04 | 0.000820 | 0.015887 |
| *RNF43* | MSI-H vs CBD | 26.74 | 0.000384 | 0.016348 |
| *ARID1A* | NSMP vs MSI-H | 0.27 | 0.001304 | 0.023140 |
| *CTCF* | MSI-H vs CBD | 16.80 | 0.000863 | 0.026270 |
| *MLL2* | MSI-H vs CBD | 16.80 | 0.000863 | 0.026270 |
| *NF1* | NSMP vs MSI-H | 0.05 | 0.001891 | 0.026846 |
| *HNF1A* | NSMP vs MSI-H | 0.05 | 0.001891 | 0.026846 |
| *MSH6* | NSMP vs MSI-H | 0.05 | 0.001891 | 0.026846 |
| *AKT2* | TP53mut vs MSI-H | 15.82 | 0.004039 | 0.029669 |
| *PAX5* | TP53mut vs MSI-H | 0.04 | 0.004989 | 0.035421 |
| *APC* | TP53mut vs MSI-H | 0.11 | 0.005603 | 0.037962 |
| *TSC1* | TP53mut vs MSI-H | 0.07 | 0.006060 | 0.037962 |
| *ATR* | TP53mut vs MSI-H | 0.07 | 0.006060 | 0.037962 |
| *FLCN* | TP53mut vs MSI-H | 0.07 | 0.006060 | 0.037962 |
| *CCNE1* | TP53mut vs NSMP | 7.57 | 0.001209 | 0.051492 |
| *EP300* | NSMP vs MSI-H | 0.05 | 0.004304 | 0.057298 |
| *ERBB2* | TP53mut vs MSI-H | 4.62 | 0.009872 | 0.060077 |
| *PIK3R1* | TP53mut vs MSI-H | 0.37 | 0.013758 | 0.081399 |

Genes significantly different (q < 0.1) between groups defined by molecular classification using a Chi-square or Fisher’s exact test.

Abbreviation: MSI-H, high microsatellite instability; TP53mut, *TP53* mutant; NSMP, no significant molecular profile; CBD, cannot be determined.

^1^Fisher's Exact Test for Count Data; Pearson's Chi-squared test

^2^Benjamini-Hochberg (BH) procedure for multiple hypothesis testing correction

**Table S4. Pairwise Comparisons of Significant Genes Between Histological Subtypes**

Pairwise comparisons by histology

| **gene** | **comparison** | **odds_ratio** | **p** | **q** |
| --- | --- | --- | --- | --- |
| *TP53* | Endometrioid vs Serous | 0.01 | 1.60e-30 | 3.41e-28 |
| *PTEN* | Endometrioid vs Serous | 28.35 | 7.71e-22 | 8.21e-20 |
| *ARID1A* | Endometrioid vs Serous | 16.61 | 6.34e-15 | 4.50e-13 |
| *TP53* | Endometrioid vs Carcinosarcoma | 0.06 | 2.91e-12 | 6.20e-10 |
| *CTCF* | Endometrioid vs Serous | 37.67 | 7.06e-09 | 3.76e-07 |
| *JAK1* | Endometrioid vs Serous | 55.23 | 7.07e-08 | 3.01e-06 |
| *PTEN* | Endometrioid vs Carcinosarcoma | 8.11 | 4.30e-08 | 4.58e-06 |
| *CTNNB1* | Endometrioid vs Serous | 29.71 | 3.45e-07 | 1.22e-05 |
| *ARID1A* | Endometrioid vs Carcinosarcoma | 8.61 | 3.82e-07 | 2.71e-05 |
| *CCNE1* | Endometrioid vs Serous | 0.07 | 3.31e-06 | 0.000101 |
| *PIK3R1* | Endometrioid vs Serous | 5.16 | 1.19e-05 | 0.000298 |
| *BCOR* | Endometrioid vs Serous | 36.50 | 1.26e-05 | 0.000298 |
| *MLL2* | Endometrioid vs Serous | 12.56 | 2.02e-05 | 0.000430 |
| *RNF43* | Endometrioid vs Serous | 34.05 | 2.59e-05 | 0.000502 |
| *PTEN* | Endometrioid vs Clear Cell | 11.47 | 3.68e-06 | 0.000784 |
| *CCNE1* | Endometrioid vs Carcinosarcoma | 0.06 | 2.71e-05 | 0.001443 |
| *TP53* | Endometrioid vs Clear Cell | 0.12 | 1.85e-05 | 0.001966 |
| *CTNNB1* | Endometrioid vs Carcinosarcoma | 29.17 | 7.44e-05 | 0.003169 |
| *PIK3CA* | Endometrioid vs Serous | 2.89 | 0.000276 | 0.004590 |
| *PPP2R1A* | Endometrioid vs Serous | 0.15 | 0.000280 | 0.004590 |
| *ERBB2* | Endometrioid vs Serous | 0.23 | 0.001053 | 0.016019 |
| *KRAS* | Endometrioid vs Serous | 3.98 | 0.001202 | 0.017066 |
| *CCND1* | Endometrioid vs Serous | 20.37 | 0.001863 | 0.022049 |
| *NFE2L2* | Endometrioid vs Serous | 20.37 | 0.001863 | 0.022049 |
| *QKI* | Endometrioid vs Serous | 20.37 | 0.001863 | 0.022049 |
| *PIK3CA* | Endometrioid vs Carcinosarcoma | 3.56 | 0.000754 | 0.026758 |
| *PTCH1* | Endometrioid vs Serous | 18.25 | 0.003767 | 0.042229 |
| *JAK1* | Endometrioid vs Carcinosarcoma | 12.56 | 0.001436 | 0.043702 |
| *AKT2* | Endometrioid vs Serous | 0.14 | 0.007476 | 0.070399 |
| *BCORL1* | Endometrioid vs Serous | 16.17 | 0.007602 | 0.070399 |
| *MAP3K1* | Endometrioid vs Serous | 16.17 | 0.007602 | 0.070399 |
| *MSH2* | Endometrioid vs Serous | 16.17 | 0.007602 | 0.070399 |
| *MYC* | Endometrioid vs Serous | 0.26 | 0.009378 | 0.083225 |
| *ASXL1* | Endometrioid vs Serous | 4.95 | 0.011533 | 0.098260 |

Genes significantly different (q < 0.1) between groups defined by histology using a Chi-square or Fisher’s exact test.

^1^Fisher's Exact Test for Count Data; Pearson's Chi-squared test

^2^Benjamini-Hochberg (BH) procedure for multiple hypothesis testing correction

**Table S5. Pairwise Comparisons of Significant Genes Between Genetic Ancestries**

Pairwise comparisons by genetic ancestry

| **gene** | **comparison** | **odds_ratio** | **p** | **q** |
| --- | --- | --- | --- | --- |
| *PTEN* | European vs African | 3.27 | 0.000230 | 0.0297 |
| *ARID1A* | European vs African | 3.79 | 0.000279 | 0.0297 |
| *KRAS* | European vs African | 5.93 | 0.001059 | 0.0722 |
| *TP53* | European vs African | 0.37 | 0.001355 | 0.0722 |

Genes significantly different (q < 0.1) between groups defined by genetic ancestry using a Chi-square or Fisher’s exact test.

^1^Fisher's Exact Test for Count Data; Pearson's Chi-squared test

^2^Benjamini-Hochberg (BH) procedure for multiple hypothesis testing correction

**Table S6. Area Deprivation Quantile Comparison**

| **Characteristic** | **Q1** N = 47*^1^* | **Q2** N = 123*^1^* | **Q3** N = 84*^1^* | **Q4+Q5** N = 27*^1^* | **p-value***^2^* |
| --- | --- | --- | --- | --- | --- |
| **Age at Diagnosis** |  |  |  |  | 0.053 |
| <50 | 2 (4.3%) | 9 (7.3%) | 4 (4.8%) | 2 (7.4%) |  |
| 50-60 | 10 (21%) | 19 (15%) | 11 (13%) | 5 (19%) |  |
| 60-70 | 16 (34%) | 57 (46%) | 42 (50%) | 4 (15%) |  |
| 70-80 | 16 (34%) | 25 (20%) | 23 (27%) | 13 (48%) |  |
| 80+ | 3 (6.4%) | 13 (11%) | 4 (4.8%) | 3 (11%) |  |
| **Race** |  |  |  |  | <0.001 |
| White | 37 (79%) | 63 (51%) | 44 (52%) | 19 (70%) |  |
| Black or African American | 3 (6.4%) | 37 (30%) | 31 (37%) | 4 (15%) |  |
| Asian | 7 (15%) | 17 (14%) | 7 (8.3%) | 2 (7.4%) |  |
| Other | 0 (0%) | 6 (4.9%) | 2 (2.4%) | 2 (7.4%) |  |
| **Ethnicity** |  |  |  |  | 0.4 |
| Not Hispanic or Latino | 43 (91%) | 105 (85%) | 71 (85%) | 20 (74%) |  |
| Hispanic or Latino | 2 (4.3%) | 10 (8.1%) | 5 (6.0%) | 5 (19%) |  |
| Unknown | 2 (4.3%) | 8 (6.5%) | 8 (9.5%) | 2 (7.4%) |  |
| **BMI Category** |  |  |  |  | 0.3 |
| Non-Obese | 28 (60%) | 61 (54%) | 37 (46%) | 15 (65%) |  |
| Class 1 Obesity | 9 (19%) | 29 (26%) | 24 (30%) | 2 (8.7%) |  |
| Class 2 Obesity | 8 (17%) | 11 (9.7%) | 11 (14%) | 2 (8.7%) |  |
| Class 3 Obesity | 2 (4.3%) | 12 (11%) | 8 (10%) | 4 (17%) |  |
| Unknown | 0 | 10 | 4 | 4 |  |
| **Predominant Genetic Ancestry** |  |  |  |  | 0.11 |
| European | 33 (70%) | 58 (47%) | 40 (48%) | 17 (63%) |  |
| African | 4 (8.5%) | 36 (29%) | 31 (37%) | 5 (19%) |  |
| South Asian | 3 (6.4%) | 12 (9.8%) | 4 (4.8%) | 2 (7.4%) |  |
| Admixed American | 1 (2.1%) | 6 (4.9%) | 3 (3.6%) | 2 (7.4%) |  |
| East Asian | 3 (6.4%) | 5 (4.1%) | 3 (3.6%) | 0 (0%) |  |
| Unknown | 3 (6.4%) | 6 (4.9%) | 3 (3.6%) | 1 (3.7%) |  |
| **Stage** |  |  |  |  | 0.9 |
| I | 21 (45%) | 49 (40%) | 31 (37%) | 10 (37%) |  |
| II | 1 (2.1%) | 3 (2.4%) | 2 (2.4%) | 0 (0%) |  |
| III | 10 (21%) | 29 (24%) | 21 (25%) | 11 (41%) |  |
| IV | 15 (32%) | 42 (34%) | 30 (36%) | 6 (22%) |  |
| **Grade** |  |  |  |  | 0.6 |
| G1 | 8 (17%) | 18 (15%) | 6 (7.1%) | 4 (15%) |  |
| G2 | 6 (13%) | 16 (13%) | 14 (17%) | 4 (15%) |  |
| G3 | 33 (70%) | 89 (72%) | 64 (76%) | 19 (70%) |  |
| **Molecular Classification** |  |  |  |  | 0.3 |
| TP53mut | 26 (58%) | 59 (53%) | 47 (62%) | 13 (50%) |  |
| NSMP | 11 (24%) | 27 (24%) | 12 (16%) | 10 (38%) |  |
| MSI-H | 8 (18%) | 26 (23%) | 17 (22%) | 3 (12%) |  |
| Unknown | 2 | 11 | 8 | 1 |  |
| **Histology** |  |  |  |  | 0.2 |
| Endometrioid | 14 (30%) | 51 (41%) | 33 (39%) | 12 (44%) |  |
| Serous | 14 (30%) | 42 (34%) | 33 (39%) | 8 (30%) |  |
| Carcinosarcoma | 14 (30%) | 21 (17%) | 7 (8.3%) | 4 (15%) |  |
| Clear Cell | 5 (11%) | 7 (5.7%) | 8 (9.5%) | 2 (7.4%) |  |
| Other | 0 (0%) | 2 (1.6%) | 3 (3.6%) | 1 (3.7%) |  |
| **TMB Category** |  |  |  |  | 0.2 |
| < 10 mutations per megabase | 41 (87%) | 92 (75%) | 68 (81%) | 24 (89%) |  |
| ≥ 10 mutations per megabase | 6 (13%) | 31 (25%) | 16 (19%) | 3 (11%) |  |
| **Insurance Type** |  |  |  |  | 0.4 |
| Public | 18 (38%) | 61 (50%) | 35 (42%) | 17 (63%) |  |
| Private | 11 (23%) | 30 (24%) | 18 (21%) | 6 (22%) |  |
| Uninsured | 0 (0%) | 1 (0.8%) | 2 (2.4%) | 0 (0%) |  |
| Other | 1 (2.1%) | 0 (0%) | 1 (1.2%) | 0 (0%) |  |
| Unknown | 17 (36%) | 31 (25%) | 28 (33%) | 4 (15%) |  |

Comparison of demographic and clinical characteristics between area deprivation index state quantiles. Quantiles 4 and 5 were grouped due to a low number of patients from these regions.

Abbreviations: Q1-4, quantiles 1-4.

^1^n (%)

^2^Fisher's Exact Test for Count Data with simulated p-value (based on 2000 replicates); Pearson's Chi-squared test

^3^Q1 denotes the least deprived state-level quantile, whereas Q5 corresponds to the most deprived

**Table S7. Multivariable Cox Proportional Hazards Model for Progression-Free Survival**

| **Parameter** |  | **Count** | **β** | **SE** | **Hazard Ratio** | **95% Wald Confidence Limits** | | **p-value** |
| --- | --- | --- | --- | --- | --- | --- | --- | --- |
| **ADI Quantile** | **Q1** | 45 | Reference | | | | | |
|  | **Q2 vs Q1** | 102 | -0.28 | 0.23 | 0.76 | 0.49 | 1.18 | 0.2217 |
|  | **Q3 vs Q1** | 72 | -0.12 | 0.25 | 0.89 | 0.55 | 1.45 | 0.6381 |
|  | **Q4+Q5 vs Q1** | 22 | 0.29 | 0.31 | 1.33 | 0.72 | 2.46 | 0.3562 |
| **Age** | **<50** | 12 | Reference | | | | | |
|  | **50-60 vs <50** | 35 | 0.72 | 0.52 | 2.05 | 0.74 | 5.72 | 0.1689 |
|  | **60-70 vs <50** | 101 | 0.77 | 0.49 | 2.17 | 0.82 | 5.72 | 0.1177 |
|  | **70-80 vs <50** | 71 | 1.05 | 0.50 | 2.87 | 1.07 | 7.70 | 0.0369* |
|  | **80+ vs <50** | 22 | 1.07 | 0.55 | 2.90 | 0.99 | 8.55 | 0.0534 |
| **FIGO 2009 Stage** | **I,II** | 97 | Reference | | | | | |
|  | **III,IV vs I,II** | 144 | 0.26 | 0.17 | 1.30 | 0.92 | 1.82 | 0.1381 |
| **Grade** | **G1** | 25 | Reference | | | | | |
|  | **G2 vs G1** | 35 | 0.36 | 0.32 | 1.44 | 0.76 | 2.72 | 0.2631 |
|  | **G3 vs G1** | 181 | 0.40 | 0.33 | 1.49 | 0.77 | 2.85 | 0.2339 |
| **Histology** | **Endometrioid** | 95 | Reference | | | | | |
|  | **Carcinosarcoma vs Endometrioid** | 41 | -0.52 | 0.31 | 0.59 | 0.32 | 1.09 | 0.0915 |
|  | **Clear Cell vs Endometrioid** | 21 | -0.35 | 0.35 | 0.71 | 0.35 | 1.42 | 0.3292 |
|  | **Serous vs Endometrioid** | 79 | -0.52 | 0.29 | 0.60 | 0.34 | 1.05 | 0.0712 |
|  | **Other vs Endometrioid** | 5 | 0.43 | 0.54 | 1.55 | 0.53 | 4.47 | 0.4225 |
| **Molecular Classification** | **TP53mut** | 141 | Reference | | | | | |
|  | **MSI-H vs TP53mut** | 48 | -1.01 | 0.27 | 0.36 | 0.21 | 0.62 | 0.0002* |
|  | **NSMP vs TP53mut** | 52 | -0.51 | 0.25 | 0.60 | 0.37 | 0.97 | 0.0377* |
| **HR Alteration Status** | **No HR Alteration** | 228 | Reference | | | | | |
|  | **HR Alteration vs. No HR Alteration** | 13 | 0.21 | 0.37 | 1.24 | 0.60 | 2.55 | 0.5684 |
| **Ancestry** | **European** | 133 | Reference | | | | | |
|  | **Admixed American vs European** | 11 | -0.12 | 0.39 | 0.89 | 0.41 | 1.90 | 0.7578 |
|  | **African vs European** | 58 | 0.65 | 0.20 | 1.91 | 1.29 | 2.83 | 0.0013* |
|  | **East Asian vs European** | 8 | 0.02 | 0.45 | 1.02 | 0.42 | 2.49 | 0.9648 |
|  | **South Asian vs European** | 20 | -0.44 | 0.31 | 0.65 | 0.35 | 1.19 | 0.1633 |
|  | **Unknown vs European** | 11 | 0.36 | 0.39 | 1.44 | 0.67 | 3.07 | 0.3526 |
| **BMI** | **Non-Obese** | 133 | Reference | | | | | |
|  | **Class 1 Obesity vs Non-Obese** | 55 | -0.49 | 0.21 | 0.61 | 0.41 | 0.92 | 0.0176* |
|  | **Class 2 Obesity vs Non-Obese** | 28 | 0.18 | 0.27 | 1.20 | 0.71 | 2.04 | 0.4948 |
|  | **Class 3 Obesity vs Non-Obese** | 25 | -0.05 | 0.27 | 0.95 | 0.56 | 1.62 | 0.8581 |

A multi-variable model controlling for ADI quantile, age at diagnosis, stage, grade, histology, molecular classification, HR alteration status, and BMI was constructed.

Abbreviations: SE, standard error; ADI, Area Deprivation Index; Q1-4, quantiles 1-4; FIGO, International Federation of Gynecology and Obstetrics; MSI-H, high microsatellite instability; TP53mut, *TP53* mutant; NSMP, no significant molecular profile; CBD, cannot be determined; BMI, body mass index.

*p-value <0.05

**Table S8. Multivariable Cox Proportional Hazards Model for Overall Survival**

| **Parameter** |  | **Count** | **β** | **SE** | **Hazard Ratio** | **95% Wald Confidence Limits** | | **p-value** |
| --- | --- | --- | --- | --- | --- | --- | --- | --- |
| **ADI Quantile** | **Q1** | 45 | Reference | | | | | |
|  | **Q2 vs Q1** | 102 | -0.29 | 0.30 | 0.75 | 0.42 | 1.34 | 0.3359 |
|  | **Q3 vs Q1** | 72 | -0.16 | 0.32 | 0.85 | 0.45 | 1.60 | 0.6122 |
|  | **Q4+Q5 vs Q1** | 22 | 0.01 | 0.40 | 1.01 | 0.46 | 2.22 | 0.9789 |
| **Age** | **<50** | 12 | Reference | | | | | |
|  | **50-60 vs <50** | 35 | 0.36 | 0.70 | 1.44 | 0.37 | 5.64 | 0.6046 |
|  | **60-70 vs <50** | 101 | 0.33 | 0.62 | 1.39 | 0.41 | 4.71 | 0.6023 |
|  | **70-80 vs <50** | 71 | 0.86 | 0.65 | 2.36 | 0.66 | 8.41 | 0.1849 |
|  | **80+ vs <50** | 22 | 1.07 | 0.72 | 2.90 | 0.71 | 11.84 | 0.1374 |
| **FIGO 2009 Stage** | **I,II** | 97 | Reference | | | | | |
|  | **III,IV vs I,II** | 144 | 0.71 | 0.23 | 2.04 | 1.30 | 3.20 | 0.0019* |
| **Grade** | **G1** | 25 | Reference | | | | | |
|  | **G2 vs G1** | 35 | 1.02 | 0.55 | 2.78 | 0.95 | 8.13 | 0.0613 |
|  | **G3 vs G1** | 181 | 0.73 | 0.59 | 2.07 | 0.65 | 6.57 | 0.2174 |
| **Histology** | **Endometrioid** | 95 | Reference | | | | | |
|  | **Carcinosarcoma vs Endometrioid** | 41 | 0.13 | 0.43 | 1.14 | 0.49 | 2.65 | 0.7647 |
|  | **Clear Cell vs Endometrioid** | 21 | -0.04 | 0.51 | 0.96 | 0.35 | 2.63 | 0.9391 |
|  | **Serous vs Endometrioid** | 79 | -0.17 | 0.40 | 0.84 | 0.39 | 1.84 | 0.6634 |
|  | **Other vs Endometrioid** | 5 | -0.02 | 0.72 | 0.98 | 0.24 | 4.06 | 0.9804 |
| **Molecular Classification** | **TP53mut** | 141 | Reference | | | | | |
|  | **MSI-H vs TP53mut** | 48 | -1.40 | 0.43 | 0.25 | 0.11 | 0.57 | 0.0010* |
|  | **NSMP vs TP53mut** | 52 | -0.79 | 0.33 | 0.45 | 0.24 | 0.86 | 0.0159* |
| **HR Alteration Status** | **No HR Alteration** | 228 | Reference | | | | | |
|  | **HR Alteration vs. No HR Alteration** | 13 | -0.35 | 0.65 | 0.71 | 0.20 | 2.52 | 0.5922 |
| **Ancestry** | **European** | 133 | Reference | | | | | |
|  | **Admixed American vs European** | 11 | -0.08 | 0.54 | 0.92 | 0.32 | 2.67 | 0.8783 |
|  | **African vs European** | 58 | 0.54 | 0.25 | 1.72 | 1.05 | 2.84 | 0.0322* |
|  | **East Asian vs European** | 8 | 0.68 | 0.53 | 1.97 | 0.69 | 5.62 | 0.2041 |
|  | **South Asian vs European** | 20 | 0.46 | 0.37 | 1.59 | 0.76 | 3.30 | 0.2190 |
|  | **Unknown vs European** | 11 | -0.09 | 0.64 | 0.91 | 0.26 | 3.16 | 0.8825 |
| **BMI** | **Non-Obese** | 133 | Reference | | | | | |
|  | **Class 1 Obesity vs Non-Obese** | 55 | -0.12 | 0.28 | 0.89 | 0.51 | 1.54 | 0.6684 |
|  | **Class 2 Obesity vs Non-Obese** | 28 | 0.59 | 0.35 | 1.80 | 0.91 | 3.57 | 0.0924 |
|  | **Class 3 Obesity vs Non-Obese** | 25 | 0.50 | 0.35 | 1.66 | 0.84 | 3.28 | 0.1485 |

A multi-variable model controlling for ADI quantile, age at diagnosis, stage, grade, histology, molecular classification, HR alteration status, and BMI was constructed.

Abbreviations: SE, standard error; ADI, Area Deprivation Index; Q1-4, quantiles 1-4; FIGO, International Federation of Gynecology and Obstetrics; MSI-H, high microsatellite instability; TP53mut, *TP53* mutant; NSMP, no significant molecular profile; CBD, cannot be determined; BMI, body mass index.

*p-value <0.05

**Table S9. Multivariable Cox Proportional Hazards Model for Progression-Free Survival – Race**

Progression-Free Survival

| **Parameter** |  | **Count** | **β** | **SE** | **Hazard Ratio** | **95% Wald Confidence Limits** | | **p-value** |
| --- | --- | --- | --- | --- | --- | --- | --- | --- |
| **ADI Quantile** | **Q1** | 45 | Reference | | | | | |
|  | **Q2 vs Q1** | 102 | -0.32 | 0.23 | 0.72 | 0.46 | 1.14 | 0.16 |
|  | **Q3 vs Q1** | 72 | -0.14 | 0.25 | 0.87 | 0.53 | 1.43 | 0.59 |
|  | **Q4+Q5 vs Q1** | 22 | 0.28 | 0.31 | 1.32 | 0.72 | 2.42 | 0.37 |
| **Age** | **<50** | 12 | Reference | | | | | |
|  | **50-60 vs <50** | 35 | 0.80 | 0.52 | 2.22 | 0.80 | 6.15 | 0.13 |
|  | **60-70 vs <50** | 101 | 0.82 | 0.50 | 2.26 | 0.86 | 5.99 | 0.10 |
|  | **70-80 vs <50** | 71 | 1.06 | 0.50 | 2.89 | 1.08 | 7.74 | 0.04* |
|  | **80+ vs <50** | 22 | 1.15 | 0.55 | 3.15 | 1.07 | 9.29 | 0.04* |
| **FIGO 2009 Stage** | **I,II** | 97 | Reference | | | | | |
|  | **III,IV vs I,II** | 144 | 0.23 | 0.17 | 1.26 | 0.90 | 1.77 | 0.18 |
| **Grade** | **G1** | 25 | Reference | | | | | |
|  | **G2 vs G1** | 35 | 0.36 | 0.32 | 1.43 | 0.76 | 2.71 | 0.26 |
|  | **G3 vs G1** | 181 | 0.43 | 0.33 | 1.54 | 0.81 | 2.95 | 0.19 |
| **Histology** | **Endometrioid** | 95 | Reference | | | | | |
|  | **Carcinosarcoma vs Endometrioid** | 41 | -0.57 | 0.31 | 0.56 | 0.30 | 1.04 | 0.07 |
|  | **Clear Cell vs Endometrioid** | 21 | -0.28 | 0.34 | 0.76 | 0.39 | 1.47 | 0.41 |
|  | **Serous vs Endometrioid** | 79 | -0.57 | 0.29 | 0.57 | 0.32 | 1.01 | 0.05* |
|  | **Other vs Endometrioid** | 5 | 0.35 | 0.53 | 1.42 | 0.50 | 4.03 | 0.51 |
| **Molecular Classification** | **TP53mut** | 141 | Reference | | | | | |
|  | **MSI-H vs TP53mut** | 48 | -1.04 | 0.27 | 0.35 | 0.21 | 0.61 | <0.01* |
|  | **NSMP vs TP53mut** | 52 | -0.57 | 0.25 | 0.57 | 0.35 | 0.92 | 0.02* |
| **HR Alteration Status** | **No HR Alteration** | 228 | Reference | | | | | |
|  | **HR Alteration vs. No HR Alteration** | 13 | 0.25 | 0.37 | 1.29 | 0.63 | 2.66 | 0.49 |
| **Race** | **White** | 146 | Reference | | | | | |
|  | **Asian vs White** | 29 | -0.30 | 0.26 | 0.74 | 0.45 | 1.24 | 0.26 |
|  | **Black or African American vs White** | 56 | 0.59 | 0.21 | 1.81 | 1.20 | 2.72 | <0.01* |
|  | **Other vs White** | 10 | 0.39 | 0.38 | 1.48 | 0.70 | 3.13 | 0.31 |
| **BMI** | **Non-Obese** | 133 | Reference | | | | | |
|  | **Class 1 Obesity vs Non-Obese** | 55 | -0.46 | 0.21 | 0.63 | 0.42 | 0.95 | 0.03* |
|  | **Class 2 Obesity vs Non-Obese** | 28 | 0.20 | 0.27 | 1.22 | 0.72 | 2.07 | 0.45 |
|  | **Class 3 Obesity vs Non-Obese** | 25 | -0.07 | 0.26 | 0.93 | 0.56 | 1.57 | 0.79 |

A multi-variable model controlling for ADI quantile, age at diagnosis, stage, grade, histology, molecular classification, HR alteration status, and BMI was constructed.

Abbreviations: SE, standard error; ADI, Area Deprivation Index; Q1-4, quantiles 1-4; FIGO, International Federation of Gynecology and Obstetrics; MSI-H, high microsatellite instability; TP53mut, *TP53* mutant; NSMP, no significant molecular profile; CBD, cannot be determined; BMI, body mass index.

*p-value <0.05

**Table S10. Multivariable Cox Proportional Hazards Model for Overall Survival – Race**

Overall Survival

| **Parameter** |  | **Count** | **β** | **SE** | **Hazard Ratio** | **95% Wald Confidence Limits** | | **p-value** |
| --- | --- | --- | --- | --- | --- | --- | --- | --- |
| **ADI Quantile** | **Q1** | 45 | Reference | | | | | |
|  | **Q2 vs Q1** | 102 | -0.23 | 0.29 | 0.80 | 0.45 | 1.42 | 0.44 |
|  | **Q3 vs Q1** | 72 | -0.10 | 0.32 | 0.91 | 0.48 | 1.70 | 0.76 |
|  | **Q4+Q5 vs Q1** | 22 | 0.05 | 0.40 | 1.05 | 0.48 | 2.31 | 0.90 |
| **Age** | **<50** | 12 | Reference | | | | | |
|  | **50-60 vs <50** | 35 | 0.25 | 0.66 | 1.28 | 0.35 | 4.64 | 0.71 |
|  | **60-70 vs <50** | 101 | 0.20 | 0.60 | 1.23 | 0.38 | 4.00 | 0.73 |
|  | **70-80 vs <50** | 71 | 0.76 | 0.61 | 2.13 | 0.64 | 7.08 | 0.22 |
|  | **80+ vs <50** | 22 | 0.95 | 0.69 | 2.59 | 0.67 | 10.06 | 0.17 |
| **FIGO 2009 Stage** | **I,II** | 97 | Reference | | | | | |
|  | **III,IV vs I,II** | 144 | 0.68 | 0.23 | 1.98 | 1.27 | 3.09 | <0.01* |
| **Grade** | **G1** | 25 | Reference | | | | | |
|  | **G2 vs G1** | 35 | 0.99 | 0.55 | 2.70 | 0.92 | 7.88 | 0.07 |
|  | **G3 vs G1** | 181 | 0.75 | 0.59 | 2.12 | 0.67 | 6.72 | 0.20 |
| **Histology** | **Endometrioid** | 95 | Reference | | | | | |
|  | **Carcinosarcoma vs Endometrioid** | 41 | 0.12 | 0.43 | 1.13 | 0.48 | 2.65 | 0.78 |
|  | **Clear Cell vs Endometrioid** | 21 | 0.00 | 0.50 | 1.00 | 0.38 | 2.64 | 1.00 |
|  | **Serous vs Endometrioid** | 79 | -0.02 | 0.72 | 0.98 | 0.24 | 4.05 | 0.98 |
|  | **Other vs Endometrioid** | 5 | -0.15 | 0.40 | 0.86 | 0.39 | 1.90 | 0.71 |
| **Molecular Classification** | **TP53mut** | 141 | Reference | | | | | |
|  | **MSI-H vs TP53mut** | 48 | -1.37 | 0.42 | 0.26 | 0.11 | 0.59 | <0.01* |
|  | **NSMP vs TP53mut** | 52 | -0.76 | 0.33 | 0.47 | 0.24 | 0.90 | 0.02* |
| **HR Alteration Status** | **No HR Alteration** | 228 | Reference | | | | | |
|  | **HR Alteration vs. No HR Alteration** | 13 | -0.40 | 0.65 | 0.67 | 0.19 | 2.38 | 0.54 |
| **Race** | **White** | 146 | Reference | | | | | |
|  | **Asian vs White** | 29 | 0.39 | 0.32 | 1.48 | 0.80 | 2.75 | 0.21 |
|  | **Black or African American vs White** | 56 | 0.43 | 0.25 | 1.54 | 0.94 | 2.54 | 0.09 |
|  | **Other vs White** | 10 | -0.49 | 0.75 | 0.61 | 0.14 | 2.64 | 0.51 |
| **BMI** | **Non-Obese** | 133 | Reference | | | | | |
|  | **Class 1 Obesity vs Non-Obese** | 55 | -0.10 | 0.28 | 0.91 | 0.53 | 1.56 | 0.73 |
|  | **Class 2 Obesity vs Non-Obese** | 28 | 0.57 | 0.35 | 1.77 | 0.89 | 3.49 | 0.10 |
|  | **Class 3 Obesity vs Non-Obese** | 25 | 0.47 | 0.34 | 1.60 | 0.82 | 3.10 | 0.17 |

A multi-variable model controlling for ADI quantile, age at diagnosis, stage, grade, histology, molecular classification, HR alteration status, and BMI was constructed.

Abbreviations: SE, standard error; ADI, Area Deprivation Index; Q1-5, quantiles 1-5; FIGO, International Federation of Gynecology and Obstetrics; MSI-H, high microsatellite instability; TP53mut, *TP53* mutant; NSMP, no significant molecular profile; CBD, cannot be determined; BMI, body mass index.

*p-value <0.05
